# Predicting Treatment-seeking Status for Alcohol Use Disorder Using Polygenic Scores and Machine Learning in a Deeply-Phenotyped Sample

**DOI:** 10.1101/2024.11.22.24317810

**Authors:** Zeal Jinwala, ReJoyce Green, Yousef Khan, Joel Gelernter, Rachel L. Kember, Emily E. Hartwell

## Abstract

**Background:** Few individuals with alcohol use disorder (AUD) receive treatment. Previous studies have shown drinking behavior, psychological problems, and substance dependence to predict treatment seeking. However, to date, no studies have incorporated polygenic scores (PGS), a measure of genetic risk for AUD.

**Methods:** Using the Yale-Penn sample, we identified 9,103 individuals diagnosed with DSM-IV AUD and indicated treatment-seeking status. We implemented a random forest (RF) model to predict treatment-seeking based on 91 clinically relevant phenotypes. We calculated AUD PGS for those with genetic data (African ancestry [AFR] n=3,192, European ancestry [EUR] n=3,553) and generated RF models for each ancestry group, first without and then with PGS. Lastly, we developed models stratified by age (< and ≥40 years old).

**Results:** 66.6% reported treatment seeking (M_age_=40.0, 62.4% male). Across models, top predictors included years of alcohol use and related psychological problems, psychiatric diagnoses, and heart disease. In the models without PGS, we found 79.8% accuracy and 0.85 AUC for EUR and 75% and 0.78 for AFR; the addition of PGS did not substantially change these metrics. PGS was the 10^th^ most important predictor for EUR and 23^rd^ for AFR. In the age-stratified analysis, PGS ranked 8^th^ for <40 and 48^th^ for ≥40 in EUR ancestry, and it ranked 14^th^ for <40 and 24^th^ for ≥40 in the AFR sample.

**Conclusion:** Alcohol use, psychiatric issues, and comorbid medical disorders were predictors of treatment seeking. Incorporating PGS did not substantially alter performance, but was a more important predictor in younger individuals with AUD.

**Highlights:**

- While alcohol use problems are common, few individuals seek treatment
- We used machine learning in a deeply-phenotyped sample to predict treatment-seeking
- We, for the first time, incorporated polygenic risk for alcohol use as a predictor
- Alcohol use variables, psychiatric issues, and medical problems were key predictors

## 1. Introduction

Alcohol use disorder (AUD), one of the most prevalent mental health disorders globally, affects more than 10% of U.S. adults (“2021 NSDUH Annual National Report | CBHSQ Data,” n.d.). AUD contributes significantly to morbidity and mortality, contributing to an estimated 20-40% of hospital admissions and 100,000 deaths annually (Li et al., 2004). The economic burden of AUD was an estimated annual U.S. cost of $185 billion in 2010 (Li et al., 2004). The etiology of AUD is complex, with genetic, neurobiological, psychological, and environmental factors contributing to the onset and maintenance of the disorder. AUD has an observed heritability of 50%, showing that genetics play an important role in the development of AUD along with environmental risk factors (Verhulst et al., 2015).

Despite the medical and societal burden of AUD, rates of treatment seeking are low. Results from the National Epidemiologic Survey on Alcohol and Related Conditions (NESARC)-III from 2012-2013 showed that 7.7% of the people with a past 12-month Diagnostic and Statistical Manual of Mental Disorders (DSM)-5 AUD diagnosis and 19.8% of people with a lifetime DSM-5 AUD diagnosis sought treatment (Grant et al., 2015). Given these low rates, it is important to understand the characteristics influencing treatment-seeking for AUD. A study comparing treatment seekers from the COMBINE study, a large, multi-center pharmacotherapy treatment trial, to non-treatment seekers from pharmacology studies showed that treatment-seeking individuals were older, had more years of education, and scored higher on drinking-related measures (e.g., consumed higher amounts of alcohol and met for more DSM symptoms) (Ray et al., 2017). The majority of these variables, in addition to others that differed between the groups, also predicted clinical outcomes in the COMBINE treatment-seeking participants, such as time to relapse and percent days abstinent.

Other studies have also highlighted how treatment-seeking individuals exhibited greater severity or impairment on numerous measures, including alcohol use, mood, and liver enzyme tests (Rohn et al., 2017). Treatment-seekers are also more likely to have comorbid mental health conditions and more severe AUD (Venegas et al., 2021). While informative, these secondary analyses of clinical research data may not reflect the general population of alcohol users due to factors such as recruitment strategies, inclusion criteria (e.g., requiring certain self-reported consumption levels), and exclusion criteria (e.g., many clinical trials exclude individuals with comorbid psychiatric diagnoses). Therefore, characterizing the differences between treatment-seekers and non-treatment-seekers in larger, more diverse samples that include genetic information and represent a wide variety of clinical presentations may inform intervention efforts for non-treatment-seeking individuals who may be at risk of AUD progression.

Previous work has used machine learning (ML) to understand factors related to AUD diagnosis, treatment outcomes, and treatment-seeking better (Hurtado et al., 2022). Within the COMBINE sample, ML was used to predict heavy alcohol use throughout AUD treatment and ML models developed showed 0.67-0.87 AUC in predicting whether a patient would return to heavy drinking during treatment (Roberts et al., 2022). Similarly, Lee et al (2019) developed a model that correctly classified 78% of AUD treatment-seeking individuals using alternating decision trees (ADTs) that utilized 10 measures (i.e., alcohol consumption, alcohol abuse diagnosis, depression, any current substance dependence, and non-psychiatric traits such as body mass index and IQ) as predictors of treatment-seeking. Though previous ML models have achieved moderate to high accuracy in classifying treatment seekers, to our knowledge no studies have yet incorporated genetic data to predict AUD treatment seeking.

Polygenic scores (PGS) are composite, weighted measures of effect sizes of individual variants across the entire genome that are associated with a given phenotype in a genome-wide association study (GWAS) and capture an individual’s risk for developing that trait. PGS are showing increasing promise as predictive tools in medicine, with current clinical applications in disciplines such as cancer and heart disease (Lambert et al., 2019). For example, the inclusion of PGS with clinical information when predicting Type II diabetes improved overall model accuracy (Hahn et al., 2022). PGS derived from alcohol-related phenotypes (e.g., alcohol consumption) have been shown to be associated with AUD diagnosis and AUD severity, indicating its potential as a useful tool for assessing risk for AUD, along with clinical measures of AUD (Lai et al., 2022). Higher PGS for alcohol dependence (AD) has been predictive of earlier ages of onset for alcohol use and AD in European (EUR) and African (AFR) ancestry individuals and a shorter latency in progression between age of onset of use and age of AD diagnosis in EUR individuals (Kranzler et al., 2023). To date though, no study has suggested that PGS yet provide enough information to have clinical utility for AUD traits.

Therefore, in the present study, we apply a random forest (RF) ML algorithm and incorporate PGS with clinically relevant phenotypic measures from a large sample of individuals with DSM-IV AUD to predict treatment-seeking status. Given the strong evidence of its association with alcohol use phenotypes and outcomes, we hypothesized that AUD PGS will be an important factor in predicting treatment seeking. We first developed a model using the whole sample to identify the top variables that predict the treatment-seeking status. We then split our model by EUR and AFR ancestry individuals to investigate PGS as an additional variable. As an exploratory analysis, we also examined the role of PGS in age-stratified models within each ancestry.

## 2. Methods

### 2.1 Sample Description

The Yale-Penn sample (N=14,040) was recruited from five academic sites in the United States for genetic studies of substance use. All sites received Institutional Review Board approval. After providing written informed consent, participants completed the Semi-Structured Assessment of Drug Dependence and Alcoholism (SSADDA), an in-depth interview (Joel Gelernter et al., 2014, 2014)comprising 24 modules providing data on demographics, environment, medical disorders, and psychiatric and substance use diagnoses and provided a sample for genotyping. Further information about recruitment and study methods can be found in (Gelernter et al., 2013; Gelernter et al., 2014; Gelernter et al., 2014; Kember et al., 2023). In this analysis, we included individuals who met the DSM-IV diagnosis AUD (alcohol dependence [AD] or alcohol abuse [AA]) and who answered at least one of the following three questions, “Have you ever brought up any problem you might have had with drinking with any professional?”, “Have you ever attended a self-help group (like Alcoholics Anonymous) for your drinking?” and “Have you ever been in a treatment program for a drinking problem?” (n=9,103; Figure 1).

**Figure 1:**
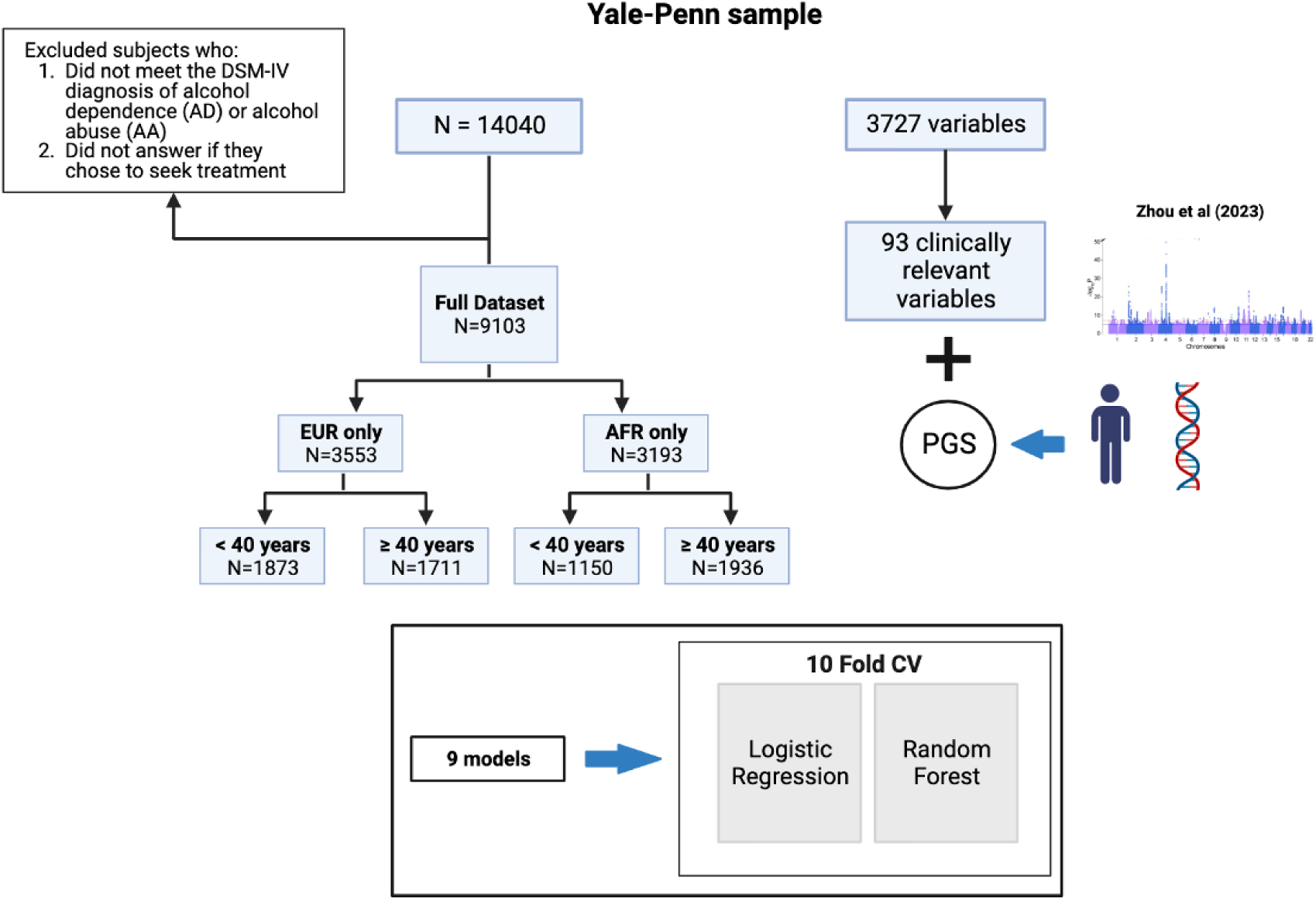
Schematic of study design

### 2.2 Genotyping, Imputation, and Polygenic Scores

Genotyping and quality control information for the Yale-Penn sample has been previously published (Gelernter et al., 2013; J. Gelernter et al., 2014; Joel Gelernter et al., 2014; Kember et al., 2023). In brief, three batches of genotyping were completed using the Illumina HumanOmni1-Quad microarray, the Illumina HumanCoreExome array, or the Illumina Multi-Ethnic Global array (Illumina, Inc.) and then imputed using the Michigan Imputation Server (Hahn et al., 2022) with 1000 Genomes phase 3 reference panel (Auton et al., 2015).

To capture genetic liability for AUD, we created ancestry-specific PGS using PRS-Continuous Shrinkage software (Ge et al., 2019) using summary statistics from a genome-wide association study of AUD in the Million Veteran Program (Zhou et al., 2023). The trait-specific effective sample sizes were used in the PGS calculation (AFR *n* = 99,583; EUR *n* = 327,814).

### 2.3 Analysis

We first tested for differences in the 91 selected variables (Supplementary Table 1), including PGS, between the treatment-seeking and non-treatment-seeking groups, and then separately for both EUR and AFR ancestries, using a chi-squared test for binary variables and a Mann-Whitney u-test for continuous variables. A Bonferroni correction was applied to account for multiple tests (p(whole sample) = 5.56 × 10^−4^, p(EUR/AFR samples) = 5.68 × 10^−4^). Missing values in the dataset were imputed using Iterative Imputation (Supplementary Table 2) and continuous variables were standardized by removing the mean and scaling to unit variance.

The dataset was split into a training sample (80%) and a test sample (20%), we then used a random forest (RF) classification model with a 10-fold repeated stratified cross-validation, distributing equal proportions of treatment-seeking and non-treatment-seeking within cross-validation split. Within each fold, we obtained the area under the curve (AUC), accuracy, precision, recall, and F1-score calculated using the test dataset. Receiver Operating Curves (ROC) and Precision-Recall Curves (PRC) were also generated within each fold (Supplementary Figure 1A-I and 2A-I). The final score is the average of performance metrics across all 10 folds. Precision indicates the fraction of positive predictions that are actually correct, while recall indicates the fraction of actually positive predictions that were predicted correctly. F1-score, the harmonic mean of precision and recall, aids in ensuring that both precision and recall are being optimized. For these three metrics, a value closer to 1 indicates good performance (Mumtaz et al., 2016). To assess which variables were important in predicting the treatment-seeking status of a participant, we employed permutation feature importance (PFI), a model-agnostic interpretability method that randomly permutes variables in the test data and obtains the drop in AUC with each permutation. PFI values were calculated within each of the 10-folds and the mean permutation values were used to rank predictors. We plotted the top twenty-five predictors for each model (Supplementary Figures 3A-I). To determine the most predictive variables, we set a PFI threshold of 1%, that is, variables that lead to a greater than 1% decrease in AUC.

We first generated a model using the entire dataset (n=9,103). We then created models including those with genetic data separately for EUR (n=3,583) and AFR (n=3,086) individuals, first without PGS and then with PGS as an additional predictor variable. Lastly, we generated age-stratified models within each ancestry, splitting by age younger than 40 (n(EUR)= 1,873, n(AFR) = 1,150) years versus 40 years and older (n(EUR)= 1,711, n(AFR) = 1,936) as this was the mean age of our sample. All analyses were performed using Python’s Scikit-learn package and the code is publicly available at (https://github.com/zealjinwala/Predicting-treatment-seeking-in-Yale-Penn-sample.git). We trained the same models described above using the logistic regression function in the Scikit-learn package for comparison with random forest. The regression coefficients and the performance metrics for logistic regression models are reported in the Supplementary materials (Supplementary Tables 3 and 4).

## 3. Results

### 3.1 Sample Description

As shown in Table 1, 9,103 individuals were included in our study, 66.6% of whom reported treatment seeking (68.6% in the EUR sample and 69.7% in the AFR sample). The average age was 40.0 ± 10.9 years (EUR = 38.6 ± 11.7, AFR = 41.8 ± 9.6). In the EUR-only subsamples, we found that PGS was higher in the treatment-seeking group than the non-treatment-seeking group (effect size= 0.58, P = 5.96 × 10^−14^) between treatment-seeking and non-treatment-seeking groups, whereas no difference was observed in the AFR-ancestry individuals. The percentage of individuals who sought treatment was higher in individuals >40 years in both ancestries. Supplementary Table 1 summarizes the baseline comparisons for all the variables included in the ML analysis. In brief, treatment-seekers in the full sample began drinking earlier, reported more years of alcohol use, were more likely to endorse craving, had blackouts, and experienced more health or emotional problems resulting from alcohol use than their non-treatment-seeking counterparts (Supplementary Table 1). Non-treatment seekers reported higher indices of socioeconomic status (e.g., higher income), while treatment-seekers reported higher rates of other substance use disorders (e.g., tobacco and cocaine dependence), medical diseases (e.g., liver disease, brain injury), and use of psychiatric medications.

**Table 1:** Demographics.

|  | **N** | **Treatment**<br>**Seeking**<br>**(%)** | **Age**<br>**(years ± SD)** | **Gender**<br>**(male, n%)** | **BMI**<br>**(mean ± SD)** |
| --- | --- | --- | --- | --- | --- |
| **Overall** | 9103 | 66.60% | 40.0 ± 10.9 | 62.40% | 28.1 ± 5.8 |
| **EUR-only** | 3583 | 68.60% | 38.57 ± 11.66 | 65.48% | 27.1 ± 5.3 |
| < 40 years | 1873 | 60.28% | 29.24 ± 5.92 | 65.08% | 26.5 ± 5.1 |
| ≥ 40 years | 1711 | 77.70% | 48.79 ± 6.85 | 65.87% | 27.8 ± 5.4 |
| **AFR-only** | 3086 | 69.70% | 41.81 ± 9.59 | 64.10% | 28.8 ± 6.0 |
| < 40 years | 1150 | 60.24% | 31.89 ± 5.68 | 59.91% | 28.9 ± 6.2 |
| ≥ 40 years | 1936 | 70.90% | 47.70 ± 5.86 | 66.58% | 28.8 ± 5.9 |

### 3.2 Classification performance of prediction models

#### 3.2.1 Whole Sample (N=9,103)

The mean cross-validation AUC and accuracy of the test dataset model generated using the complete dataset were 0.826 and 77.6%, respectively, indicating a good model fit. We report precision and recall at a threshold of 0.5. The model achieved 0.792 precision, indicating that among those predicted to seek treatment, ∼79% did indeed seek treatment. Recall was 0.904 for the same model, which is the fraction of individuals who sought treatment and were also predicted as treatment-seekers. The F-1 score for this model was 0.764, indicating that the model is effectively identifying positive cases while minimizing false positives and false negatives. The top predictor was heart disease (3.92% decrease in AUC), followed by the number of years of alcohol use, the sum of emotional and psychological problems due to drinking, and health problems due to smoking (Figure 2; Supplementary Figure 3A).

**Figure 2:**
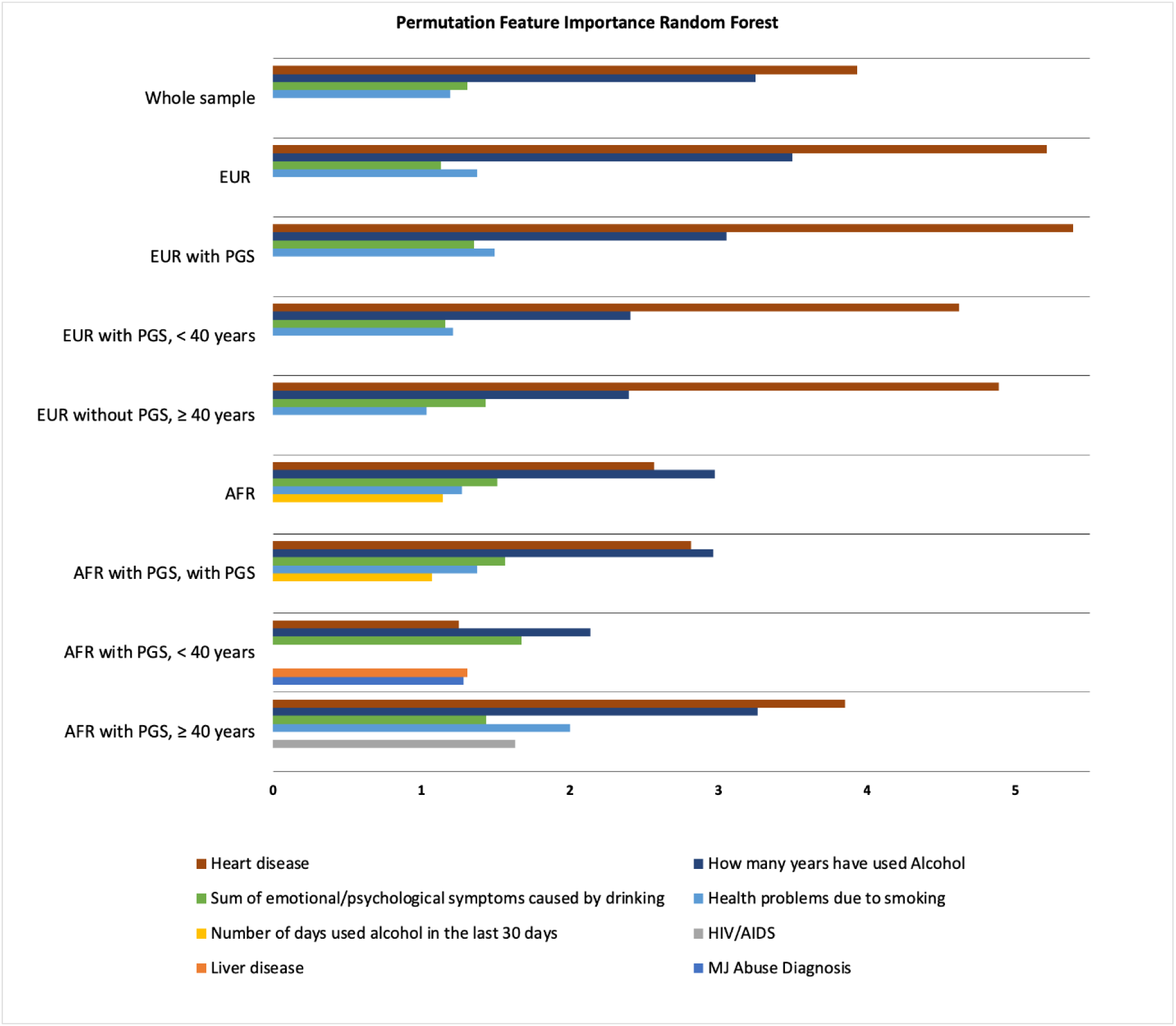
Variables with a greater than 1% permutation feature importance value in random forest models

#### 3.2.2 European ancestry (N= 3,583)

The AUC and accuracy for the models generated for the EUR dataset only were 0.850 and 79.6%, respectively. The model achieved a precision of 0.810, a recall of 0.918, and an F1-score of 0.785, indicating a good model fit. The top predictor was the number of years of alcohol use (4.94% decrease in AUC), followed by high blood pressure, the sum of emotional and psychological problems due to drinking, and the number of days of alcohol use in the last 30 days (Figure 2; Supplementary Figure 3B). The addition of PGS did not substantially improve the model performance, with an AUC of 0.847, an accuracy of 79.7%, precision, recall, and F1-score of 0.811, 0.918, and 0.786, respectively. The top predictor in this model was heart disease (3.92% decrease in AUC), followed by the number of years of alcohol use, the sum of emotional and psychological problems caused by drinking, and health problems due to smoking (Figure 2; Supplementary Figure 3C). Although it did not meet our PFI threshold, PGS was the 12^th^ most important variable.

The model AUC, prediction accuracy, and other performance metrics for models generated with EUR individuals <40 years or ≥40 years are reported in Table 2. Heart disease was the most important predictor in both models (<40 years: 4.62% decrease in AUC; ≥40 years: 4.89% decrease in AUC), followed by the number of years of alcohol use, the sum of emotional/psychological symptoms caused by drinking, and health problems due to smoking (Figure 2; Supplementary Figures 3D and E). Although causing a less than 1% decrease in AUC, PGS was the 8^th^ most important variable in the under 40 model, whereas PGS fell at 48^th^ importance in individuals 40 years and older.

**Table 2:** Performance metrics calculated on the test data.

|  | **AUC** | **Accuracy** | **Precision** | **Recall** | **F1-score** |
| --- | --- | --- | --- | --- | --- |
| **Overall** | 0.826 ± 0.015 | 0.776 ± 0.012 | 0.792 ± 0.011 | 0.904 ± 0.011 | 0.764 ± 0.014 |
| **EUR only** |  |  |  |  |  |
| without PGS | 0.850 ± 0.017 | 0.796 ± 0.023 | 0.810 ± 0.015 | 0.918 ± 0.024 | 0.785 ± 0.024 |
| with PGS | 0.847 ± 0.022 | 0.797 ± 0.019 | 0.811 ± 0.014 | 0.918 ± 0.025 | 0.786 ± 0.020 |
| with PGS, < 40 years | 0.826 ± 0.014 | 0.755 ± 0.026 | 0.764 ± 0.024 | 0.861 ± 0.025 | 0.749 ± 0.028 |
| with PGS, ≥ 40 years | 0.830 ± 0.043 | 0.835 ± 0.022 | 0.850 ± 0.018 | 0.956 ± 0.017 | 0.816 ± 0.026 |
| **AFR only** |  |  |  |  |  |
| without PGS | 0.787 ± 0.022 | 0.755 ± 0.019 | 0.774 ± 0.014 | 0.917 ± 0.017 | 0.732 ± 0.022 |
| with PGS | 0.784 ± 0.022 | 0.766 ± 0.018 | 0.781 ± 0.013 | 0.924 ± 0.021 | 0.744 ± 0.020 |
| with PGS, < 40 years | 0.762 ± 0.050 | 0.727 ± 0.043 | 0.735 ± 0.035 | 0.861 ± 0.038 | 0.716 ± 0.046 |
| with PGS, ≥ 40 years | 0.779 ± 0.032 | 0.782 ± 0.017 | 0.793 ± 0.013 | 0.960 ± 0.016 | 0.741 ± 0.023 |

#### 3.2.3 African ancestry (N=3,086)

The AFR models had an AUC of 0.787 and an accuracy of 77.4%. The precision, recall, and F1 score were 0.774, 0.917, and 0.732 respectively, suggesting that the model performed well. The top predictors were the number of years of alcohol use (2.97% decrease in AUC), heart disease, the sum of emotional and psychological problems caused by drinking, health problems due to smoking, and the number of days of alcohol use in the last 30 days (Figure 2; Supplementary Figure 3F). With PGS as an additional variable, the model performance did not change substantially, and the top predictors remained the same (Supplementary Figure 3G). PGS was the 23^rd^ most important variable with a permutation importance value of less than a 1% drop in the AUC.

The most important predictors in the <40 and ≥40 models were the number of years of alcohol use (2.14% decrease in AUC) and heart disease (3.83% decrease in AUC), respectively (Supplementary Figure 3H and I). Interestingly, liver disease and marijuana abuse diagnosis were also important predictors in the below 40 model along with the sum of the sum of emotional/psychological symptoms caused by drinking. In the above 40 model, the number of years of alcohol use, health problems due to smoking, HIV/AIDs, and the sum of emotional/psychological symptoms caused by drinking were also important predictors. PGS was the 14^th^ most important variable in the RF for individuals <40 versus 24^th^ in the model for those ≥40.

## 4. Discussion

In the present study, we used ML to predict the treatment-seeking status of individuals with a DSM-IV diagnosis of AUD in the Yale-Penn sample. In addition to assessing non-genetic predictors, we incorporated PGS as a variable in our ancestry-specific models to investigate its predictive performance for treatment-seeking. To our knowledge, this is the first study to include PGS as a variable in an ML model for predicting treatment-seeking. All models achieved an AUC greater than 0.75, indicative of good performance in distinguishing treatment-seeking and non-treatment-seeking participants. Comparable to the test accuracy of 78% achieved by Lee et al. (2019), our models achieved comparable test accuracy in the range of 72.7% to 83.5%.

In the whole sample, top predictors were heart disease, years of alcohol use, sum of emotional and psychological problems caused by drinking, and health problems due to smoking. In the EUR model, years of alcohol use, high blood pressure, the sum of emotional and psychological problems due to drinking, and the number of days of alcohol use in the past month were the top predictors. When PGS was added, it ranked 12^th,^ and the top predictors were the same as the whole sample model. In the age-stratified analysis, the top predictors were the same as the EUR with PGS model for both analyses, although PGS ranked 8^th^ for the <40 group compared to 48^th^ in the ≥40 group.

In the AFR model, the top predictors were number of years of alcohol use, heart disease, the sum of emotional and psychological problems caused by drinking, health problems due to smoking, and the number of days of alcohol use in the past month. When PGS was added, the top predictors were the same and PGS ranked 23^rd^. In the age-stratified analysis, both heart disease and years of alcohol use continued to be top predictors, however, in the under 40 group, cannabis abuse diagnosis, and liver disease were also significant whereas HIV status and health problems due to smoking were key for those over 40. PGS ranked 14^th^ in the <40 group and 24^th^ in the ≥40 group.

The inclusion of PGS as a feature did not improve predictive performance. It was among the top twenty-five features for EUR and AFR models but did not meet our set PFI threshold of 1%, indicating that variables capturing information on drinking behavior were more important for prediction. Despite the similar number of EUR and AFR participants in Yale-Penn, PGS ranked lower in the AFR group, likely due to the difference in power of the discovery of GWAS. In the age-stratified models, PGS had a higher feature importance value in models trained with participants below the age of 40, suggesting that PGS may be more useful in evaluating treatment seeking in the early course of AUD. For instance, for a patient who exhibits symptoms in early adulthood but does not seek treatment for AUD, PGS could be a useful tool in assessing their risk for disease progression and incentivizing lifestyle modification.

Along with measures of substance use, a unique feature of the Yale-Penn sample compared to electronic health record (EHR)-based biobanks is the in-depth assessment of demographic, psychological, and environmental domains. Previous studies of treatment-seeking for AD identified several measures to characterize differences between treatment-seeking and non-treatment-seeking individuals. Lee et al. (2019) reported that quality and quantity of drinking, depression, drinking-related psychological problems, and substance dependence were the measures that best predicted participants as treatment-seeking. Other studies examining the differences between non-treatment- and treatment-seeking individuals with AD also reported elevated mood and anxiety-related problems in treatment seekers (Ray et al., 2017; Rohn et al., 2017). In a sample tenfold that of previous studies, we identified variables important in predicting treatment-seeking, based on a PFI threshold of 1%. Consistent with observations from prior studies (Lee et al., 2019; Ray et al., 2017; Rohn et al., 2017), the number of years of alcohol use, a measure of drinking behavior, was one of the top predictive variables in all models, underscoring the relationship between increased AD symptoms and treatment seeking (Haass-Koffler et al., 2020; Ray et al., 2017). The medical and psychological heterogeneity of AD is also reflected by “heart disease” and “the sum of emotional and psychological symptoms due to drinking” being the other lead predictors of treatment-seeking across all models. Thus, our findings provide further validation to key predictors identified in prior studies and assess their importance relative to that of PGS. A recent genetic Mendelian randomization study established a non-linear association of increased risk of hypertension and coronary artery disease at different levels of habitual alcohol consumption (Biddinger et al., 2022), which is also evident in our results showing heart disease as a key factor that may be driving individuals to seek treatment for AUD. All variables that met the PFI threshold were positively associated with treatment-seeking in the logistic regression model, except the number of days of alcohol use in the last 30 days (Supplementary Table 4). This is possibly due to individuals who opted for treatment cutting back, recovering, or quitting drinking due to anticipation of treatment enrollment, however, it is beyond the capability of our data to investigate.

Our results should be interpreted in light of several limitations. First, the clinical measures incorporated in this study rely on self-reported interview responses and hence are subject to recall bias and underreporting. Second, the Yale-Penn sample is a cross-sectional purpose-recruited dataset, lacking longitudinal measures of disease development and treatment outcomes that may be more widely available in datasets originating from EHRs. Third, the GWAS summary statistics used to generate PGS in this study are based on data from the Million Veteran Program, an EHR-based sample that is largely male, older, and with complex comorbidities. The power of PGS also depends on the sample size of the originating GWAS, which is substantially smaller for AFR ancestry. Fourth, we considered only a single alcohol-related trait. Other alcohol use traits, such as drinks per week, problematic alcohol use, or maximum habitual alcohol use, could plausibly be differentially informative for the present application. Lastly, the clinical utility of our ML approach should be assessed in light of possible false positives and false negative results. In practice, misclassifying a patient as not qualifying for seeking treatment when it is needed could delay intervention and contribute to disease progression.

In conclusion, this study aimed to evaluate a machine learning approach with the inclusion of polygenic scores in predicting treatment seeking for AUD. Our findings suggest that, in the Yale-Penn sample, where comprehensive measures capturing the complex etiology of substance use are available, the information contributed by AUD PGS is less important, particularly in individuals over 40 years of age. In settings where detailed information about diagnostic criteria, family history, and history of alcohol use is limited, such as in primary care clinics or early stages of disease onset, PGS may serve as an important measure in predicting treatment-seeking. Our study uniquely explores the role of genetic liability in treatment utilization for AUD. A similar approach replicated in EHR-derived biobank data, incorporating measures from clinical notes, pharmacy records, genetic data, and laboratory tests could further elucidate key drivers of treatment-seeking and aid in developing strategies for improving treatment utilization. We conclude further that PGS for this purpose is far from clinical utility.

## Author Contributions

EEH, ZL, and RLK conceived the study. JG acquired the data. ZL and YK curated the data. ZL conducted the analysis under the supervision of RG. ZL, YK, and EEH wrote the first draft of the manuscript. All authors reviewed and edited the manuscript.

## Data Statement

Yale-Penn data are available through dbGAP (study accession numbers phs000425.v1.p1, phs000952.v1.p1, phs000277.v2.p1).

