## Supplementary Figures for "Predicting Treatment-seeking Status for Alcohol Use Disorder Using Polygenic Scores and Machine Learning in a Deeply-Phenotyped Sample"

**Supplementary Figure 1: ROC curves of** **RF (left) and LR (right) for each dataset**

a. Whole sample

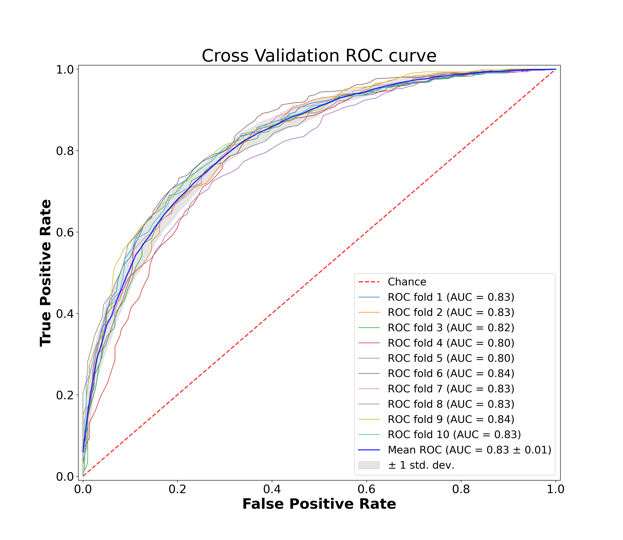

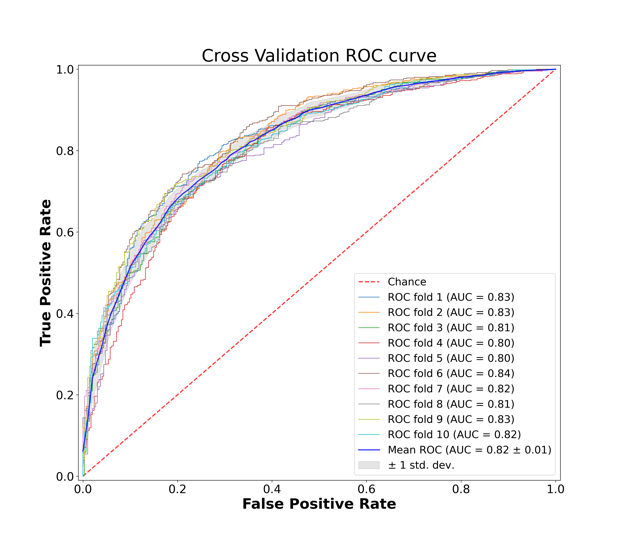

b. EUR-only without PGS

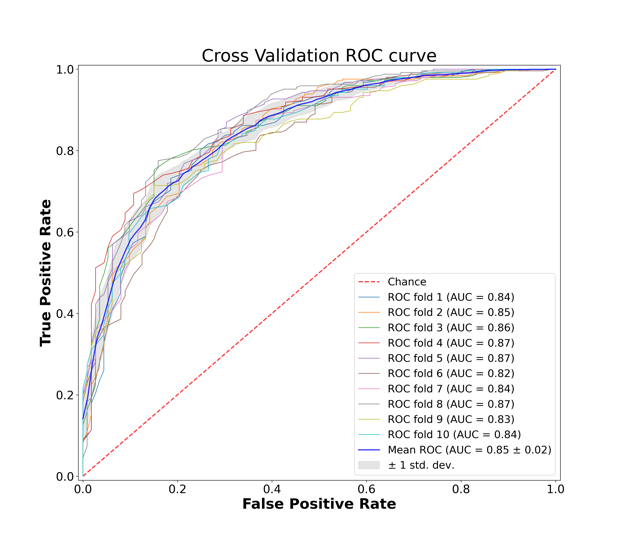

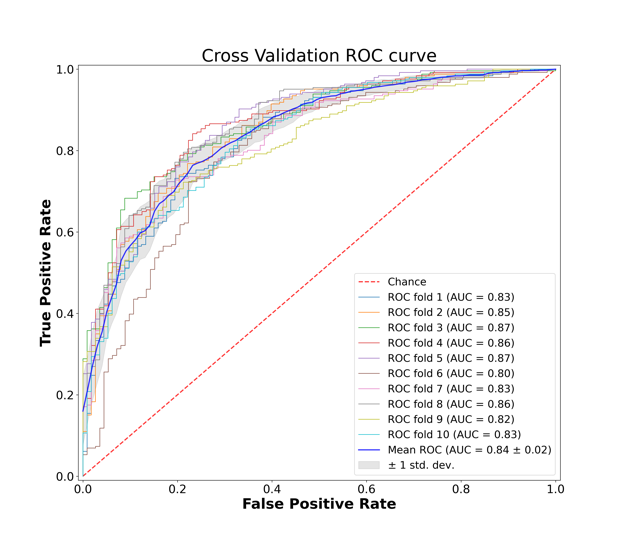

c. EUR-only with PGS

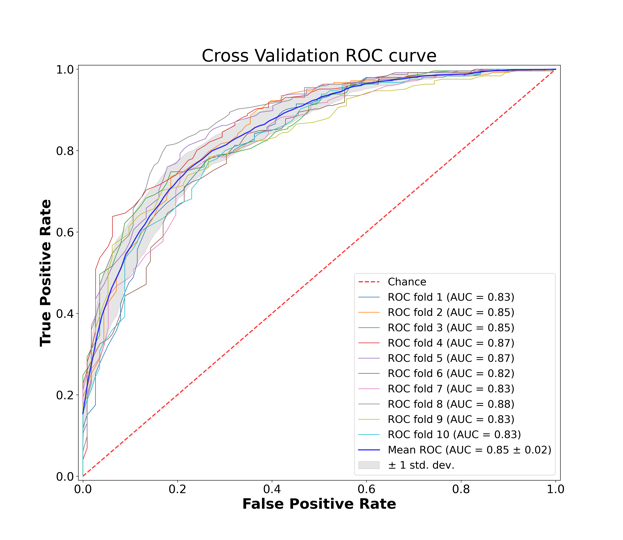

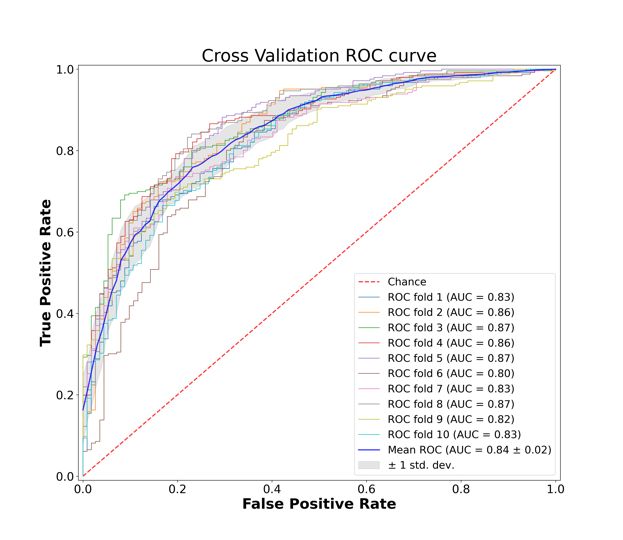

d. EUR-only with PGS, < 40 years

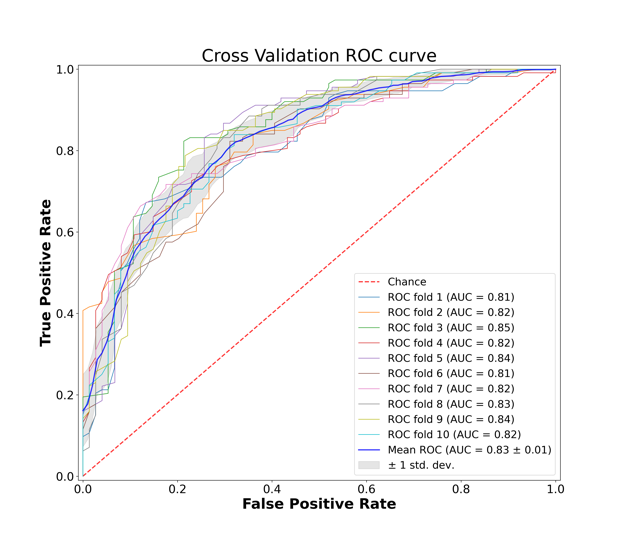

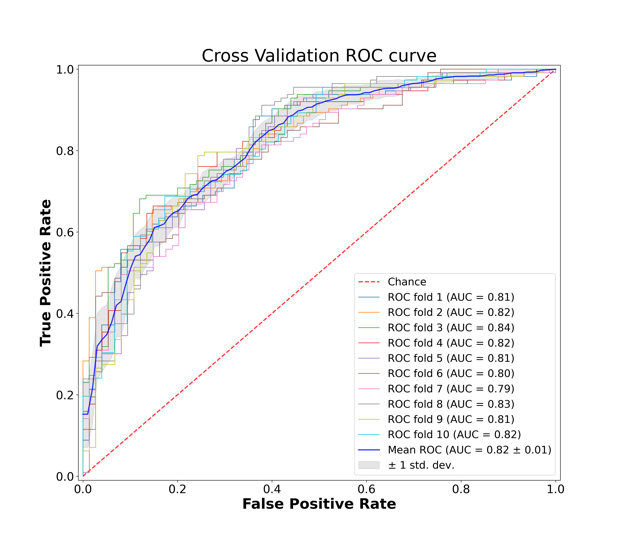

e. EUR-only with PGS, >= 40 years

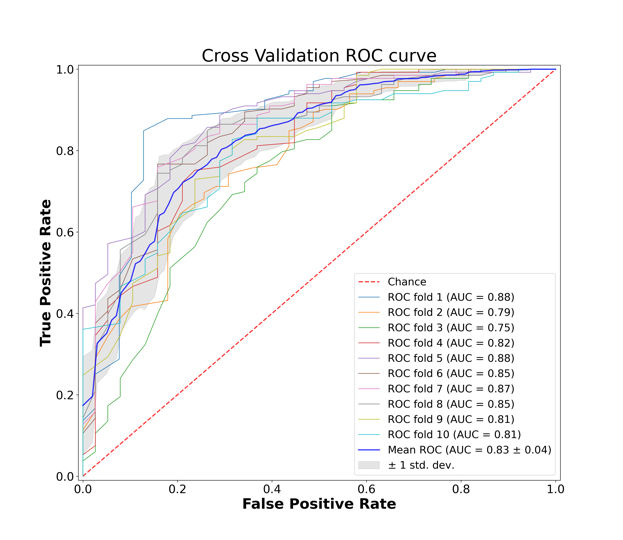

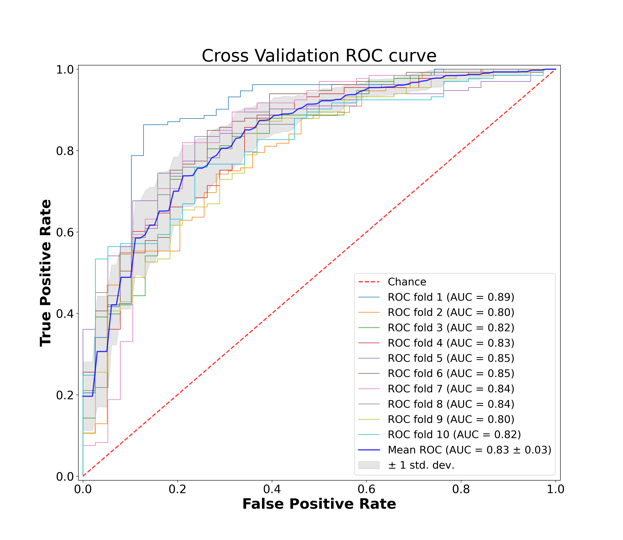

f. AFR-only without PGS

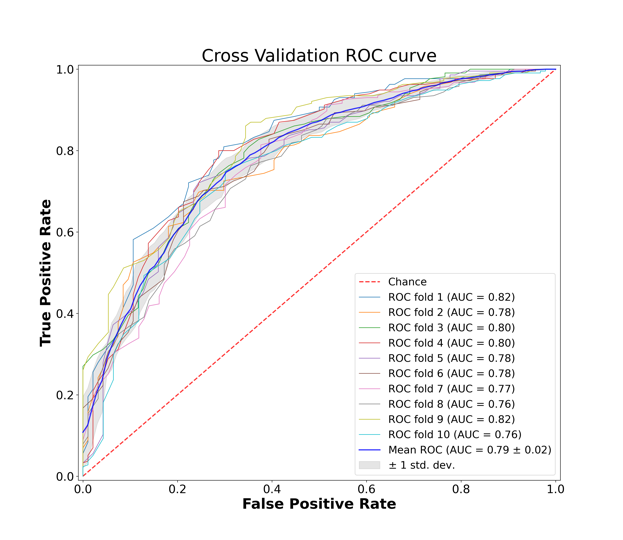

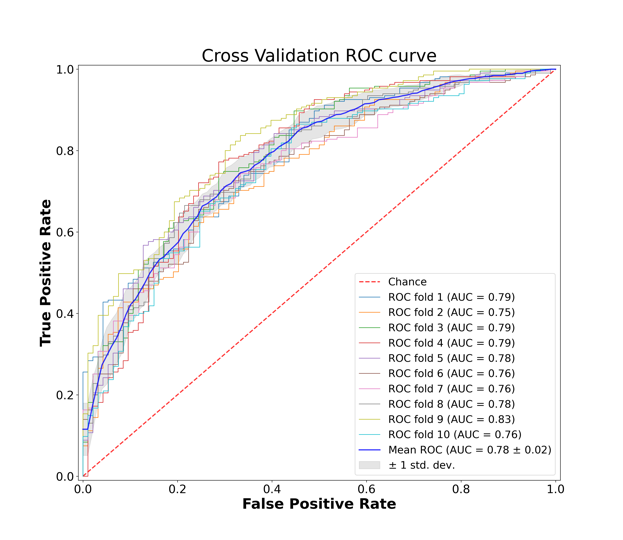

g. AFR-only with PGS

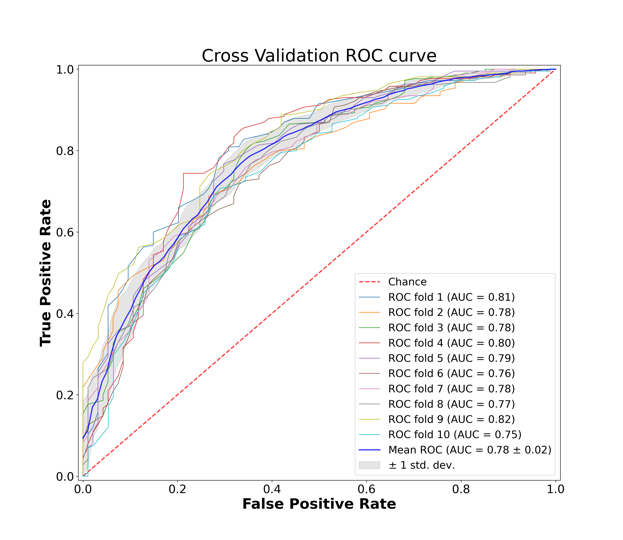

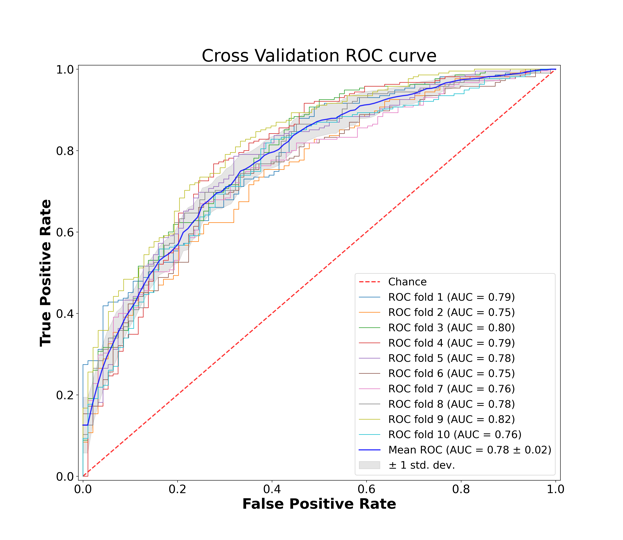

h. AFR-only with PGS, < 40 years

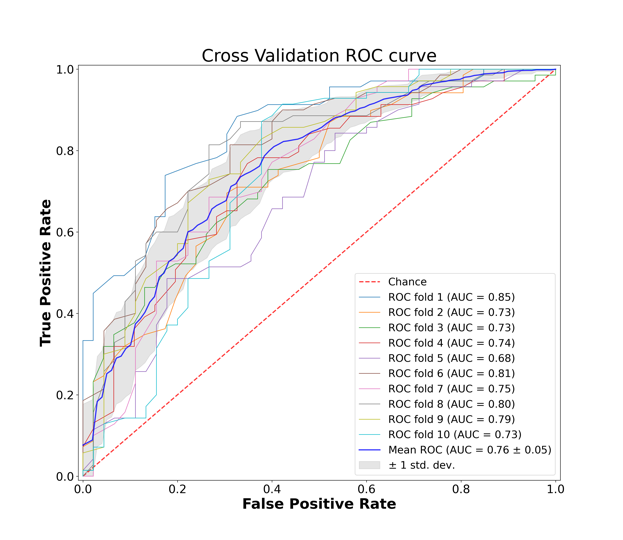

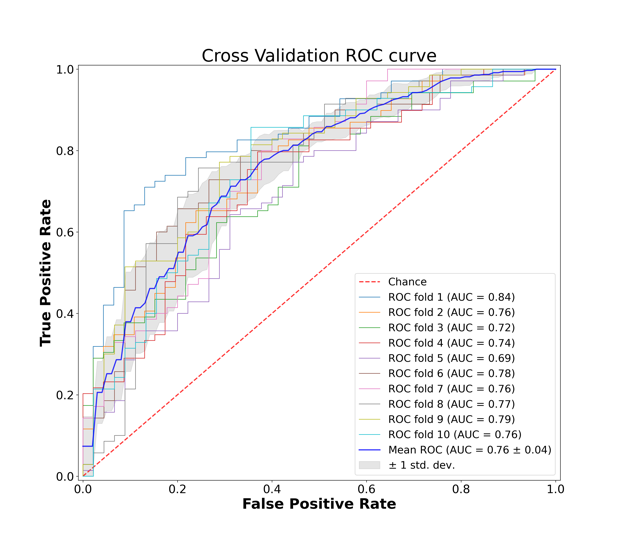

i. AFR-only with PGS, >= 40 years

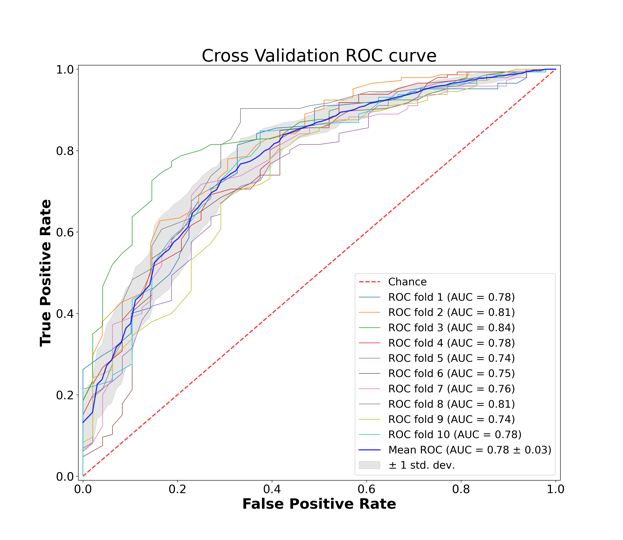
3
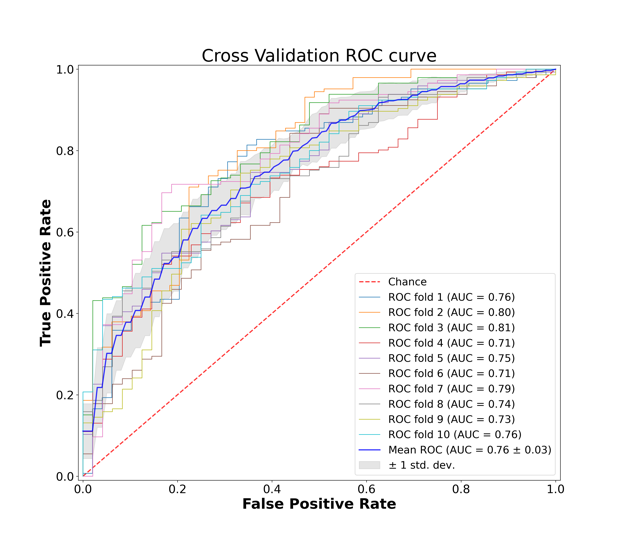

**Supplementary Figure 2: Precision-Recall curves of RF (left) and LR (right) for each dataset**

a. Whole sample

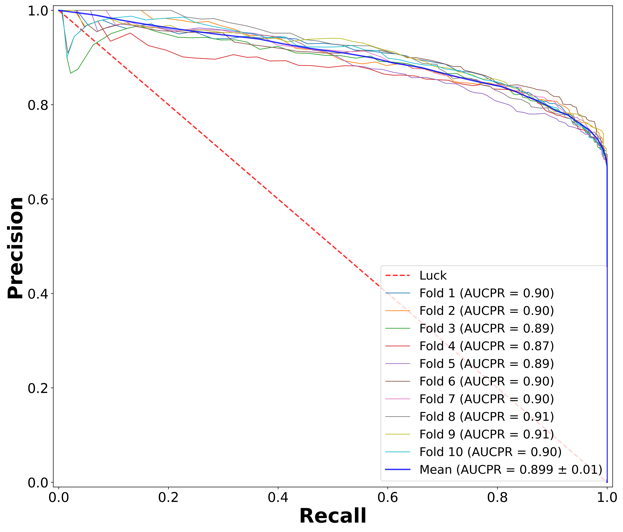

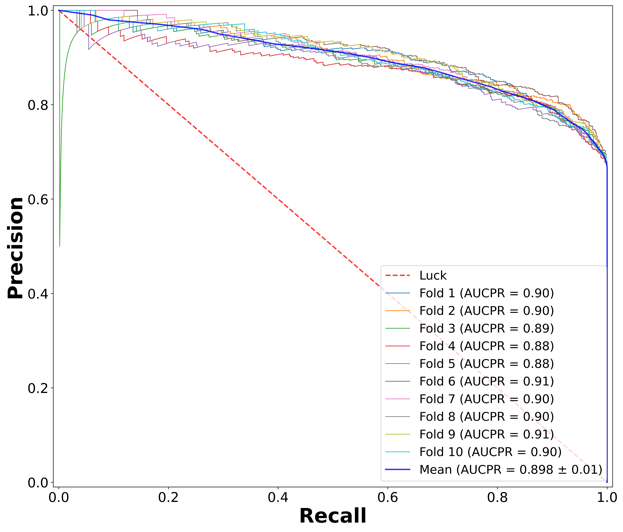

b. EUR-only without PGS

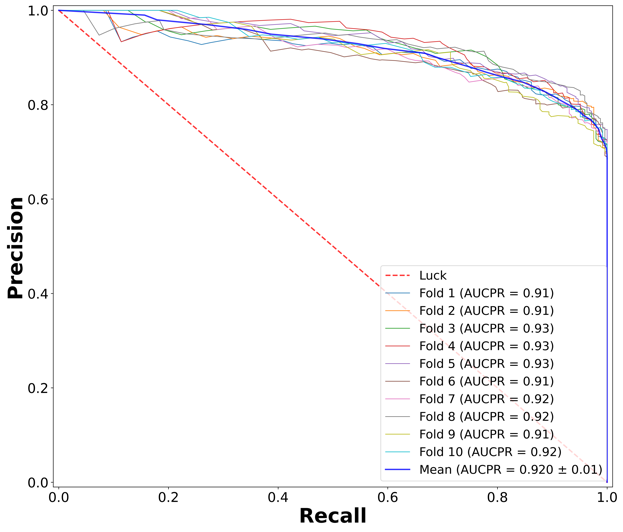

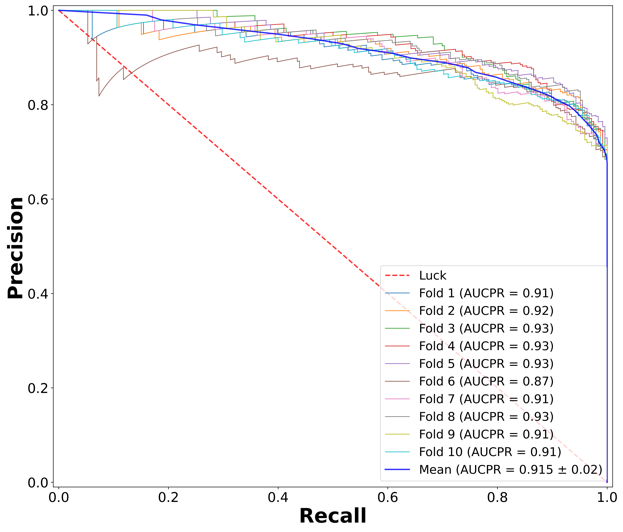

c. EUR-only with PGS

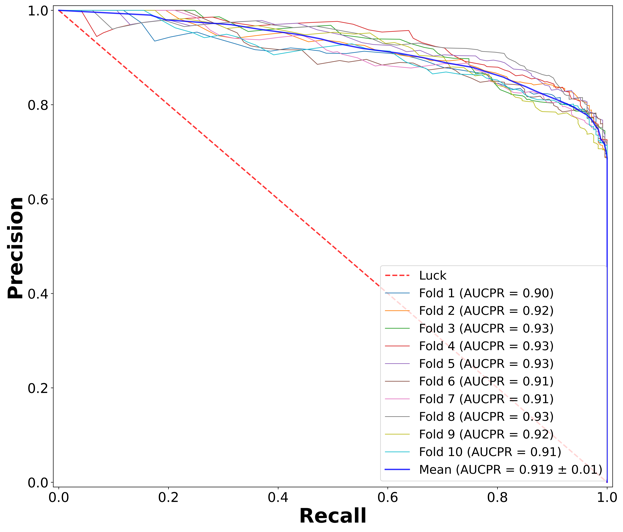

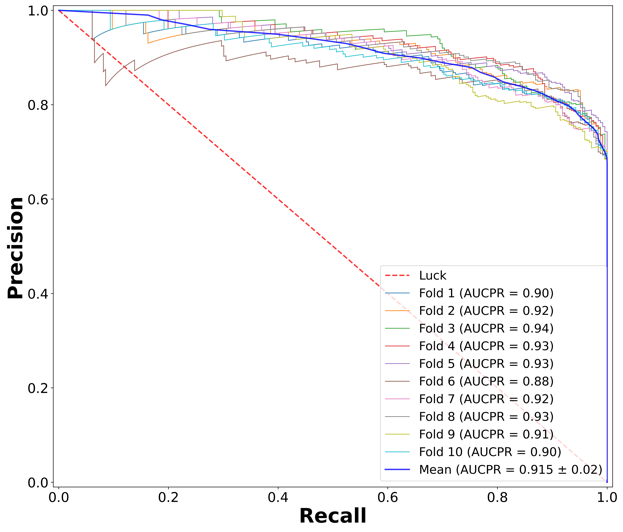

d. EUR-only with PGS, < 40 years

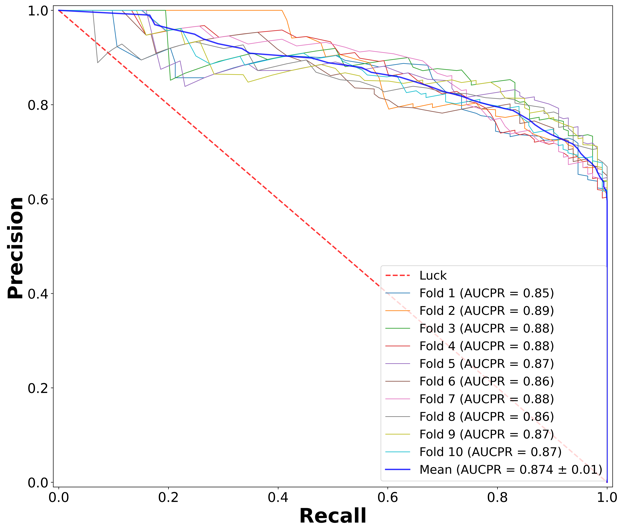

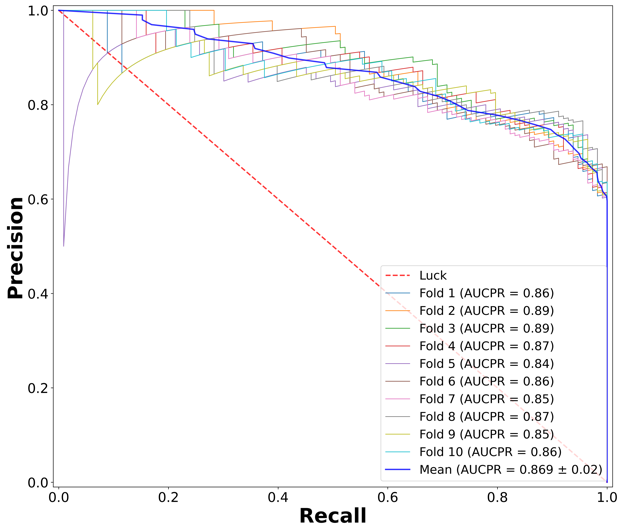

e. EUR-only with PGS, >= 40 years

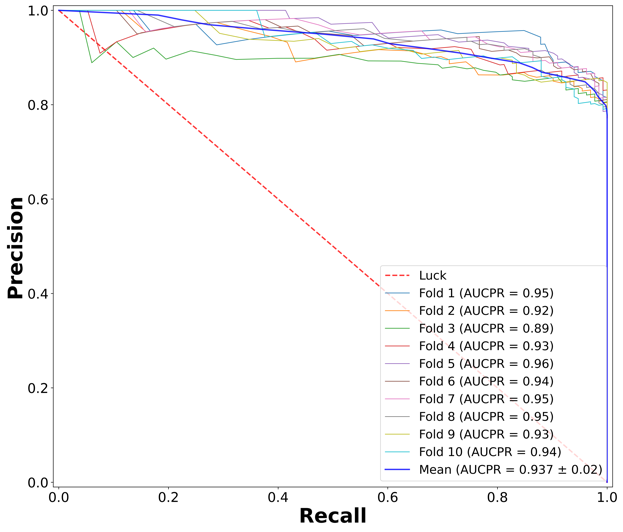

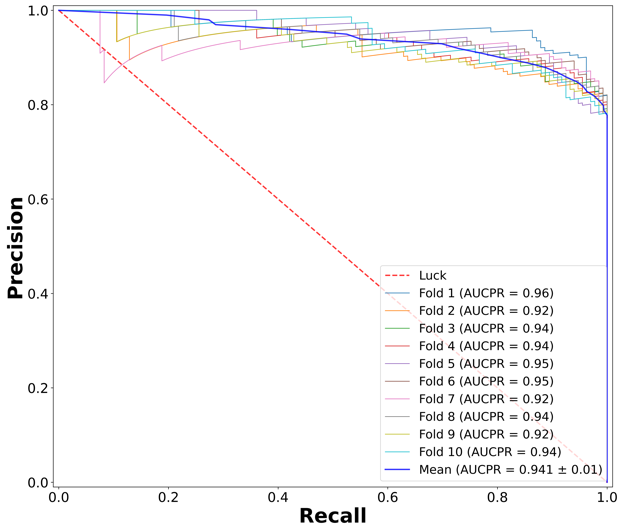

f. AFR-only without PGS

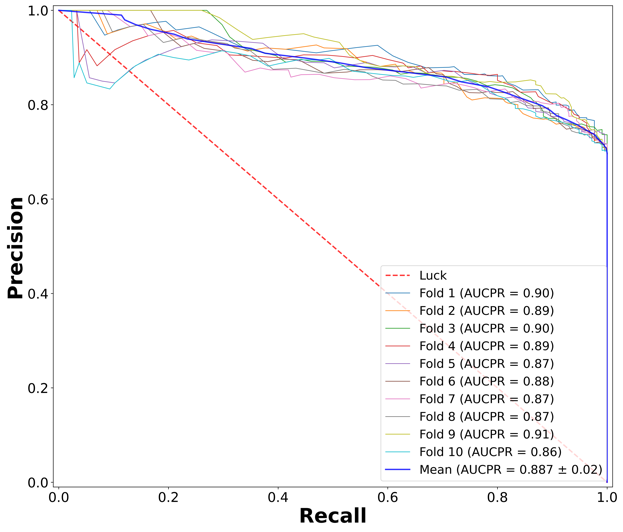

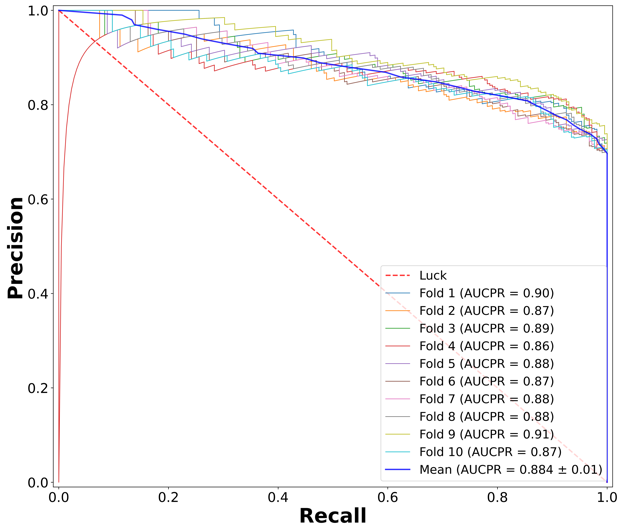

g. AFR-only with PGS

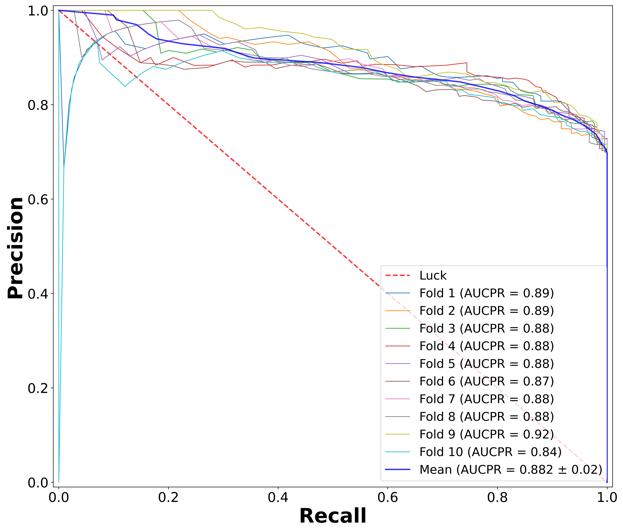

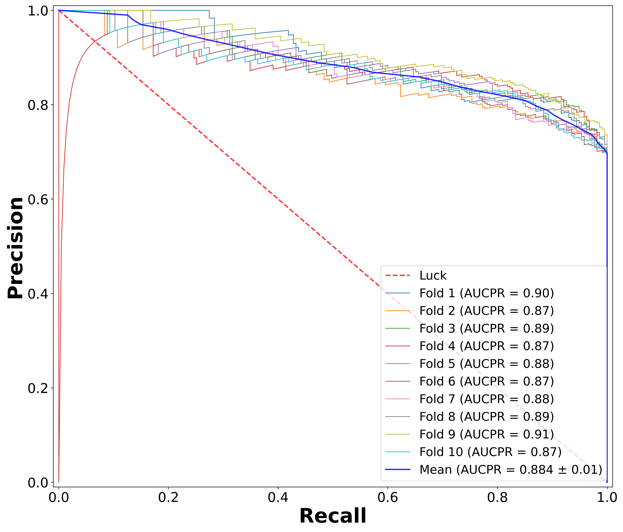

h. AFR-only with PGS, < 40 years

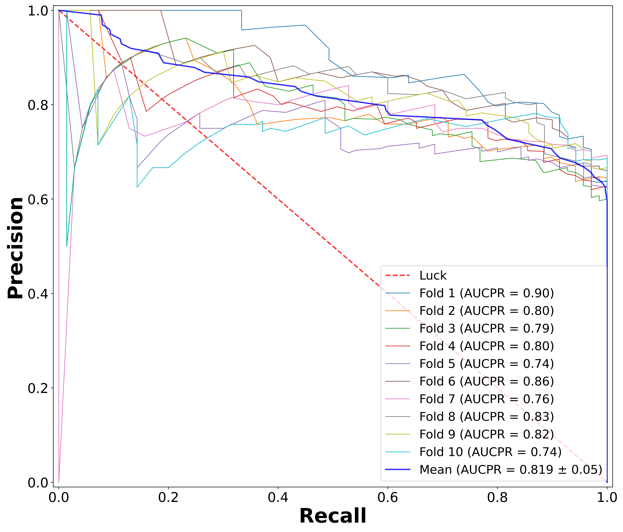

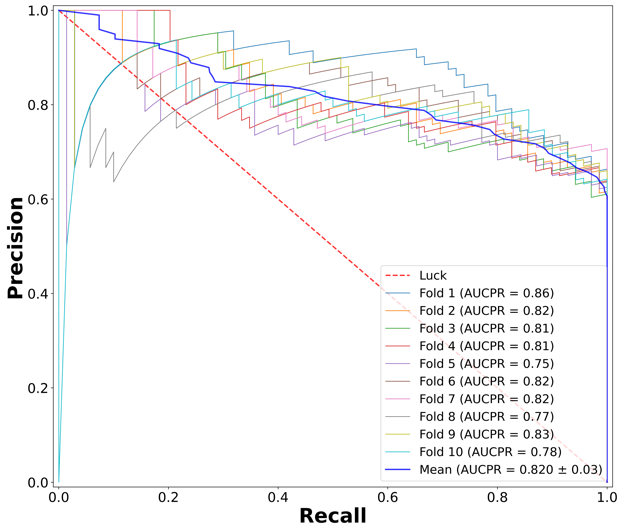

i. AFR-only with PGS, >= 40 years

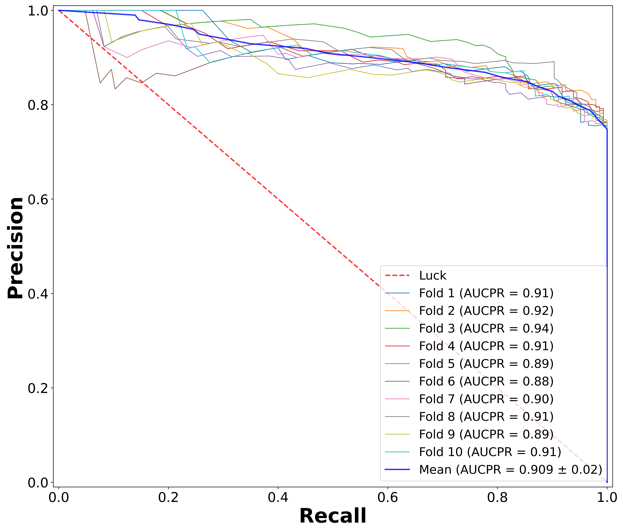

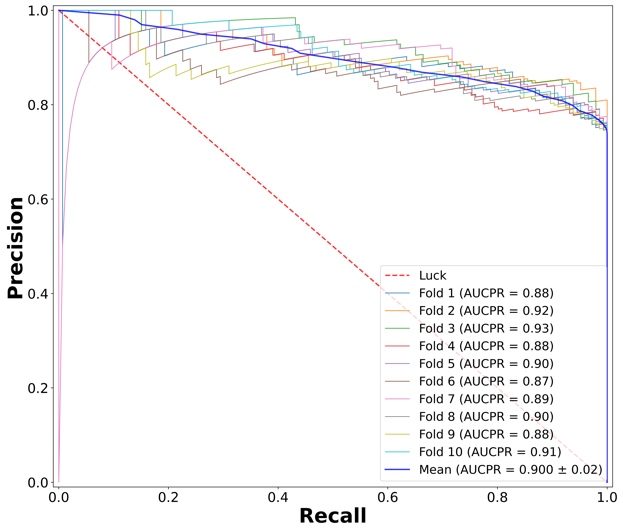

**Supplementary Figure 3: Feature Importance plots (RF and LR)**

A) Whole sample

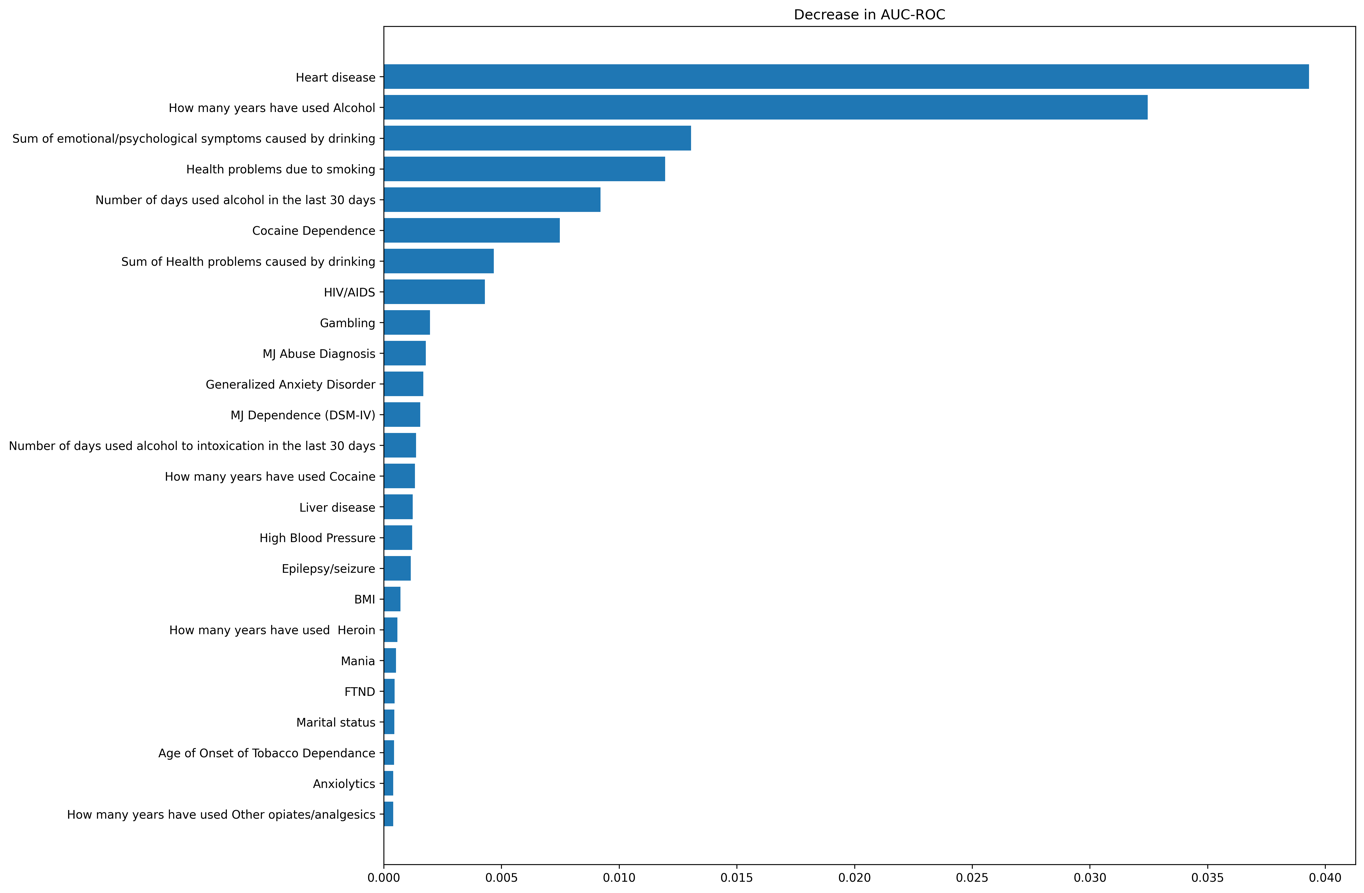

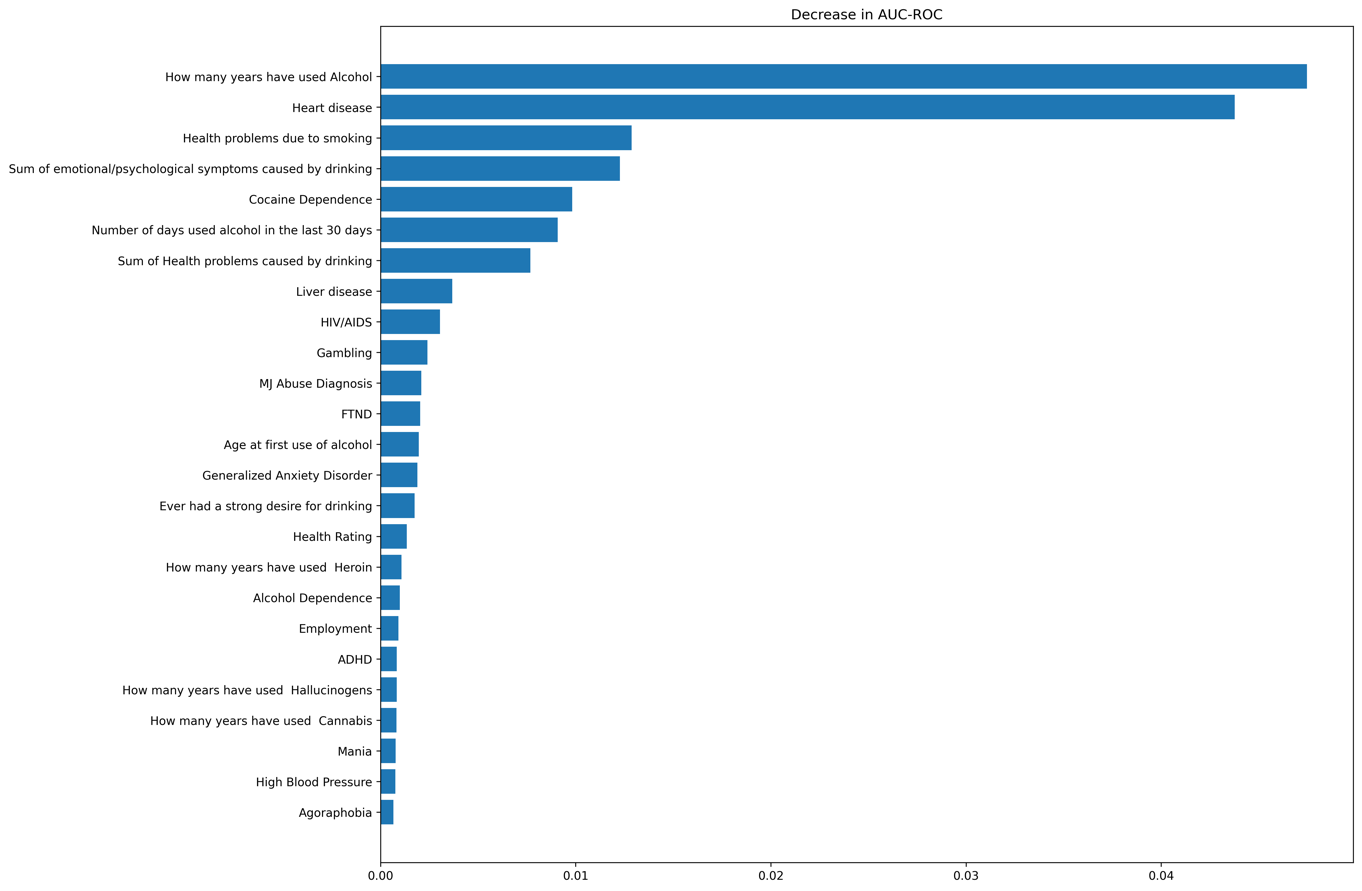

B) EUR-only without PGS

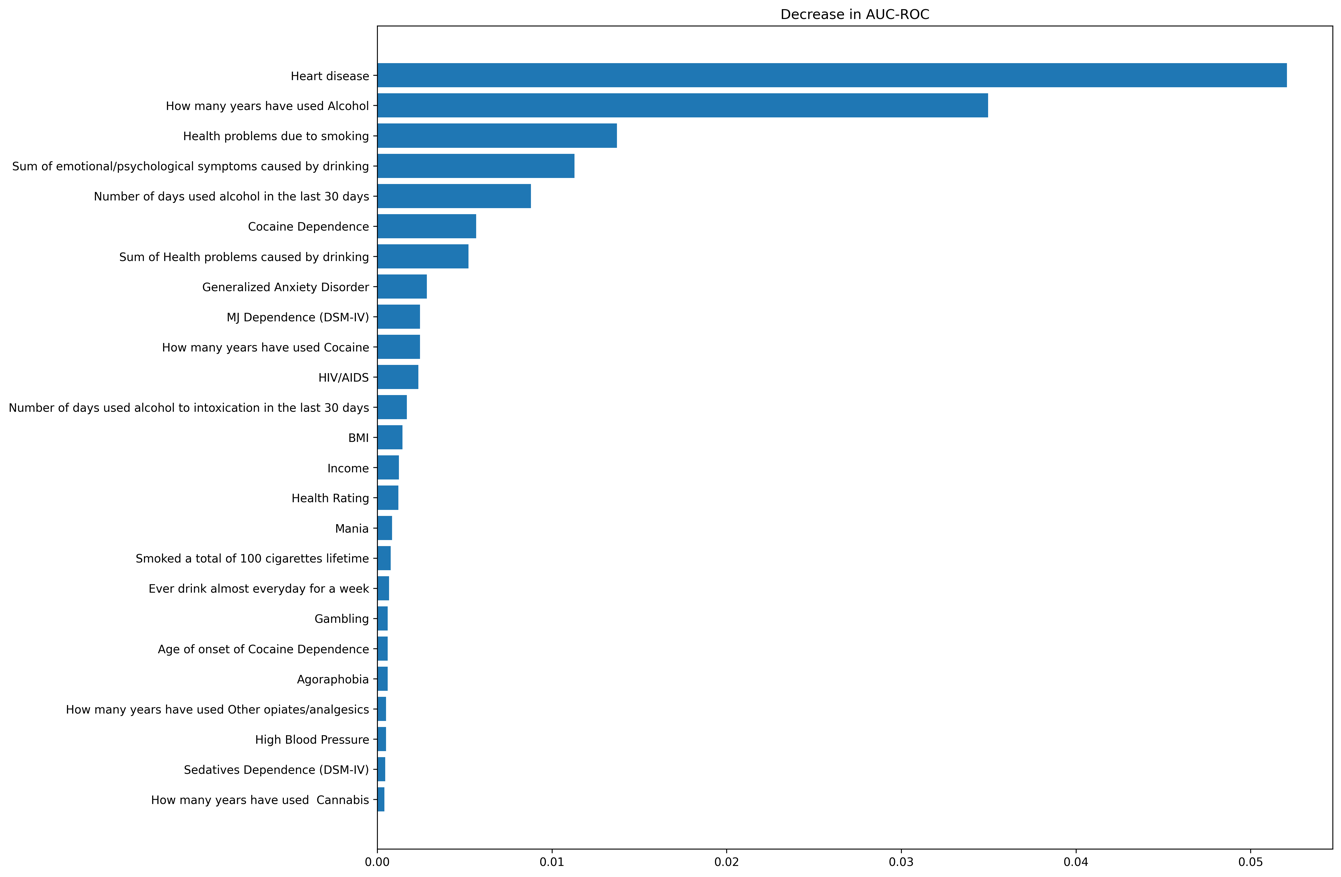

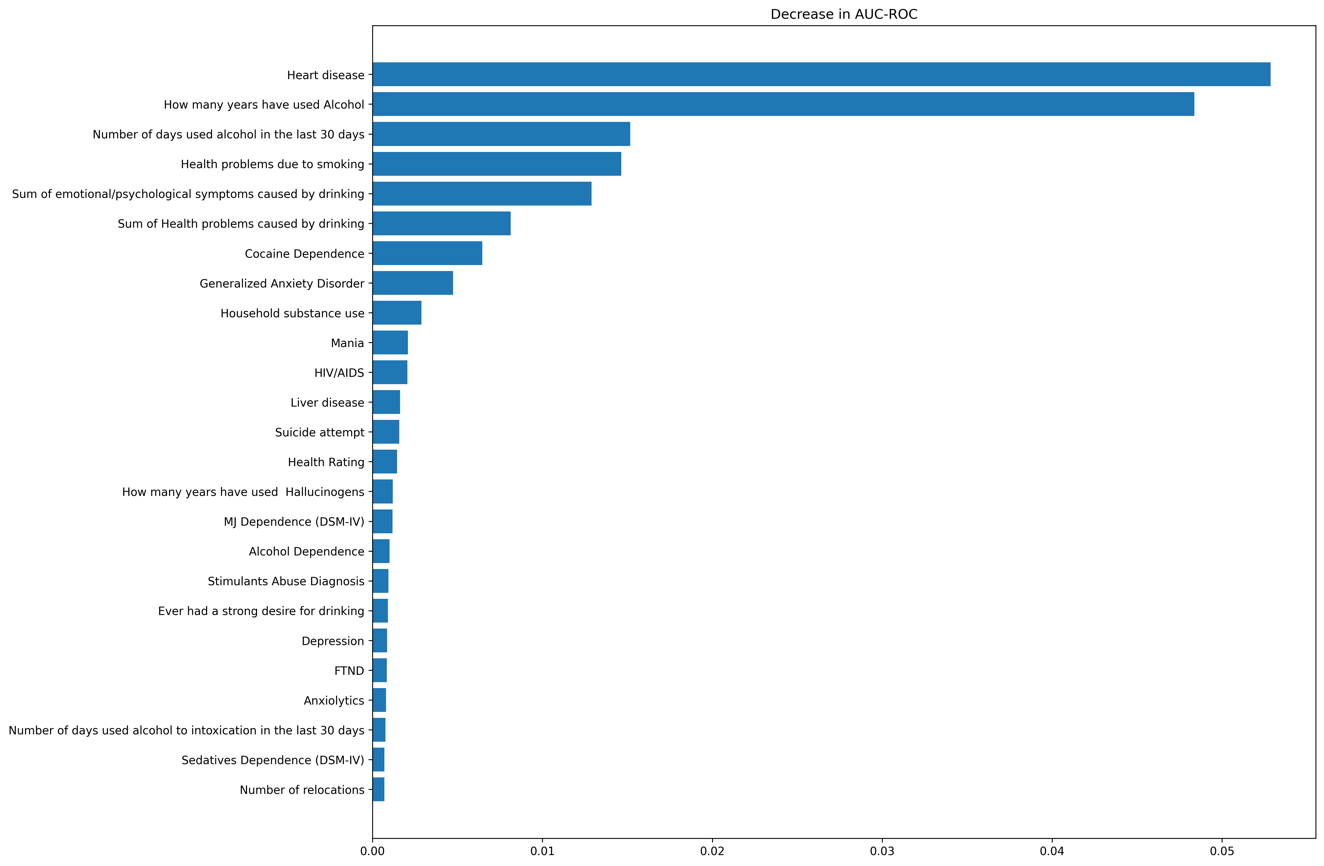

C) EUR-only with PGS

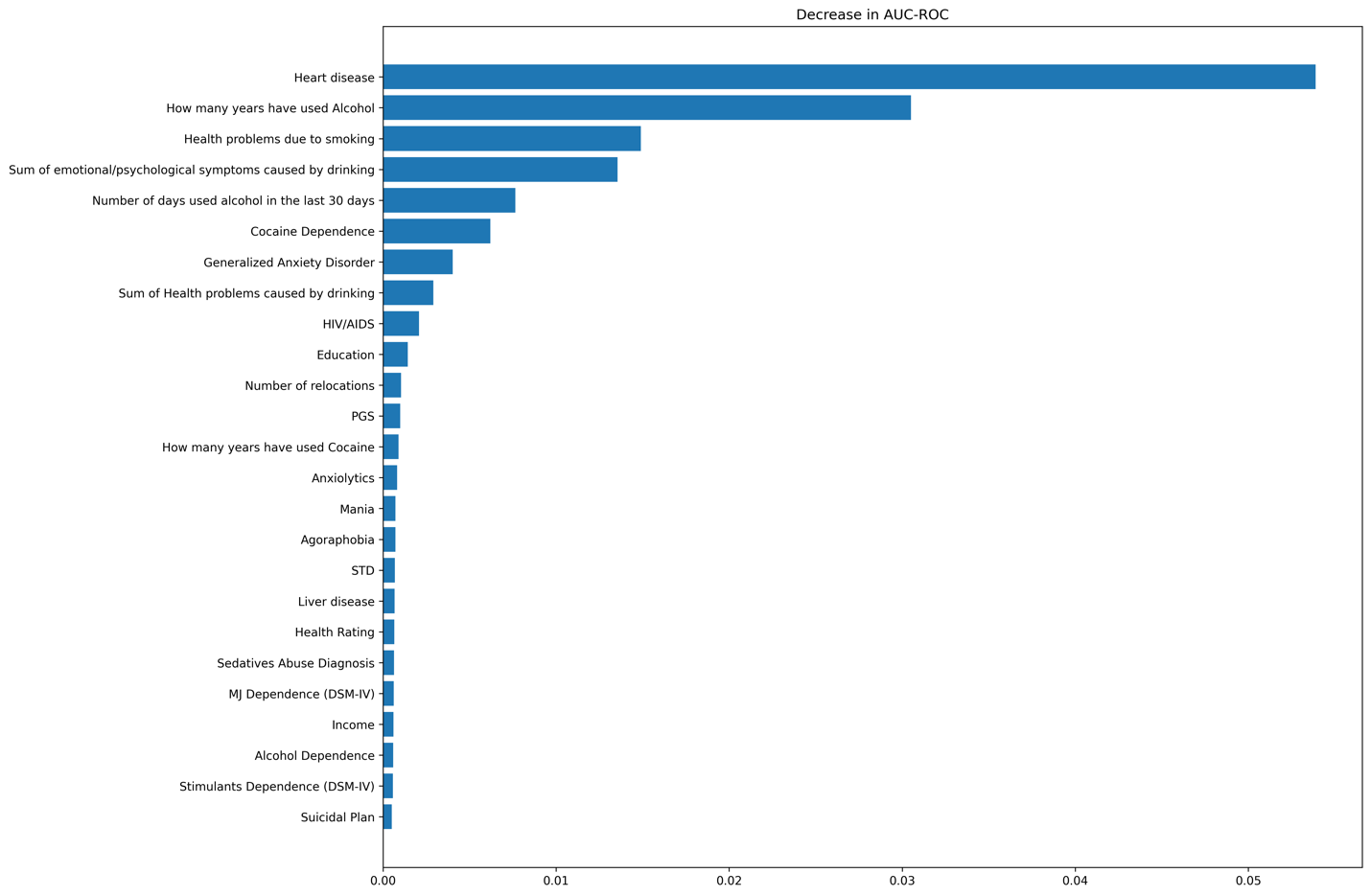

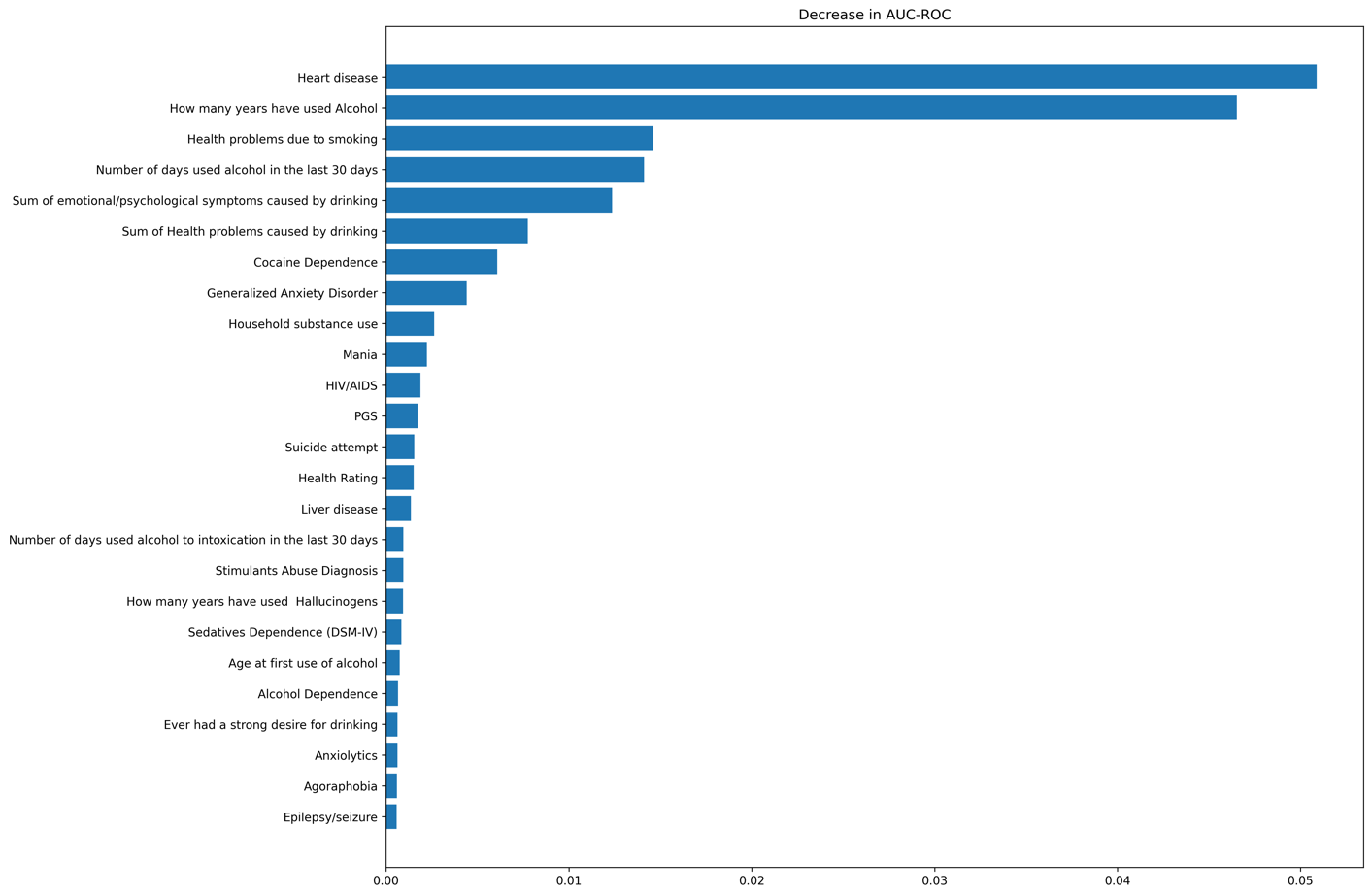

D) EUR-only with PGS, < 40 years

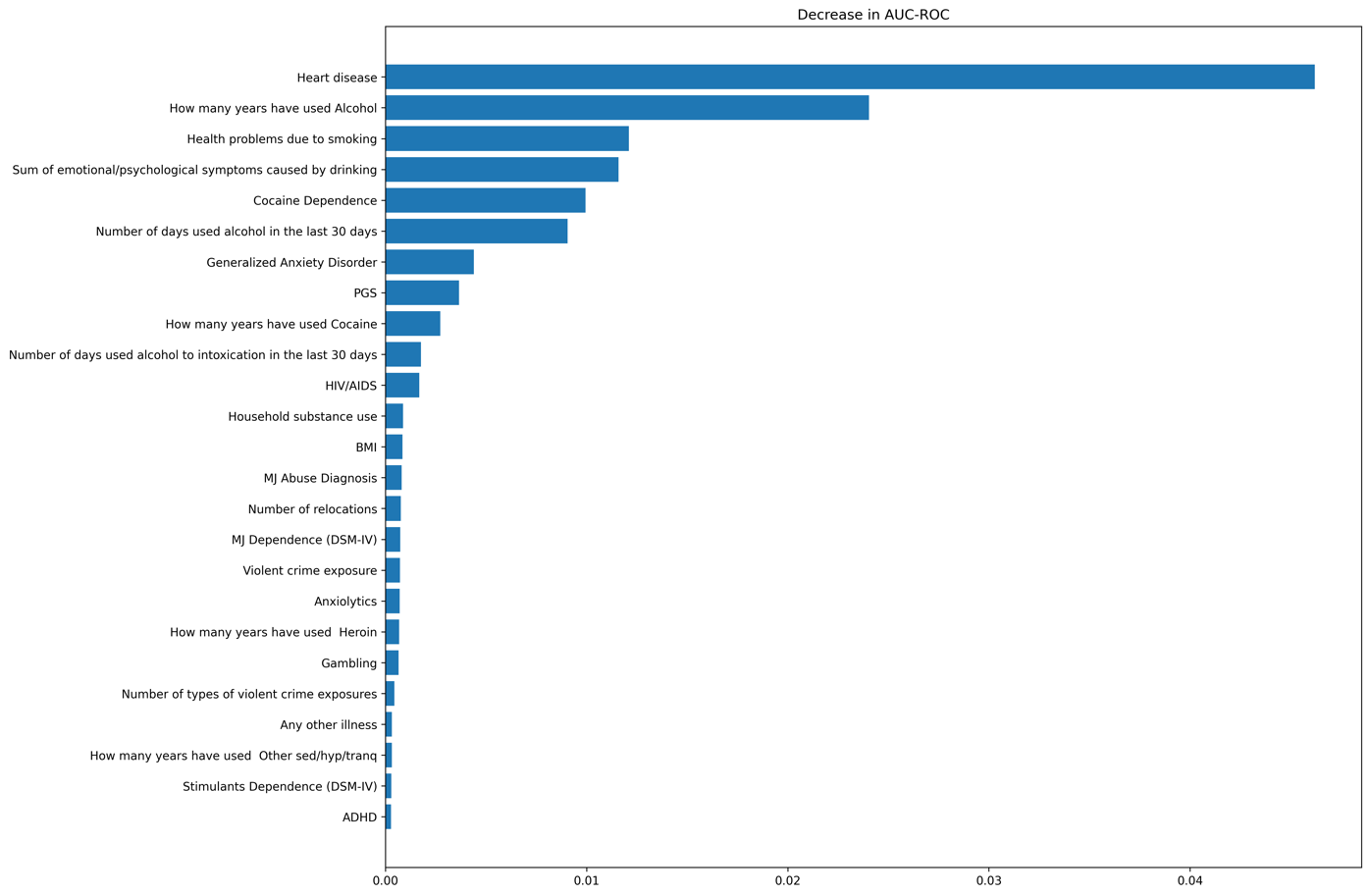

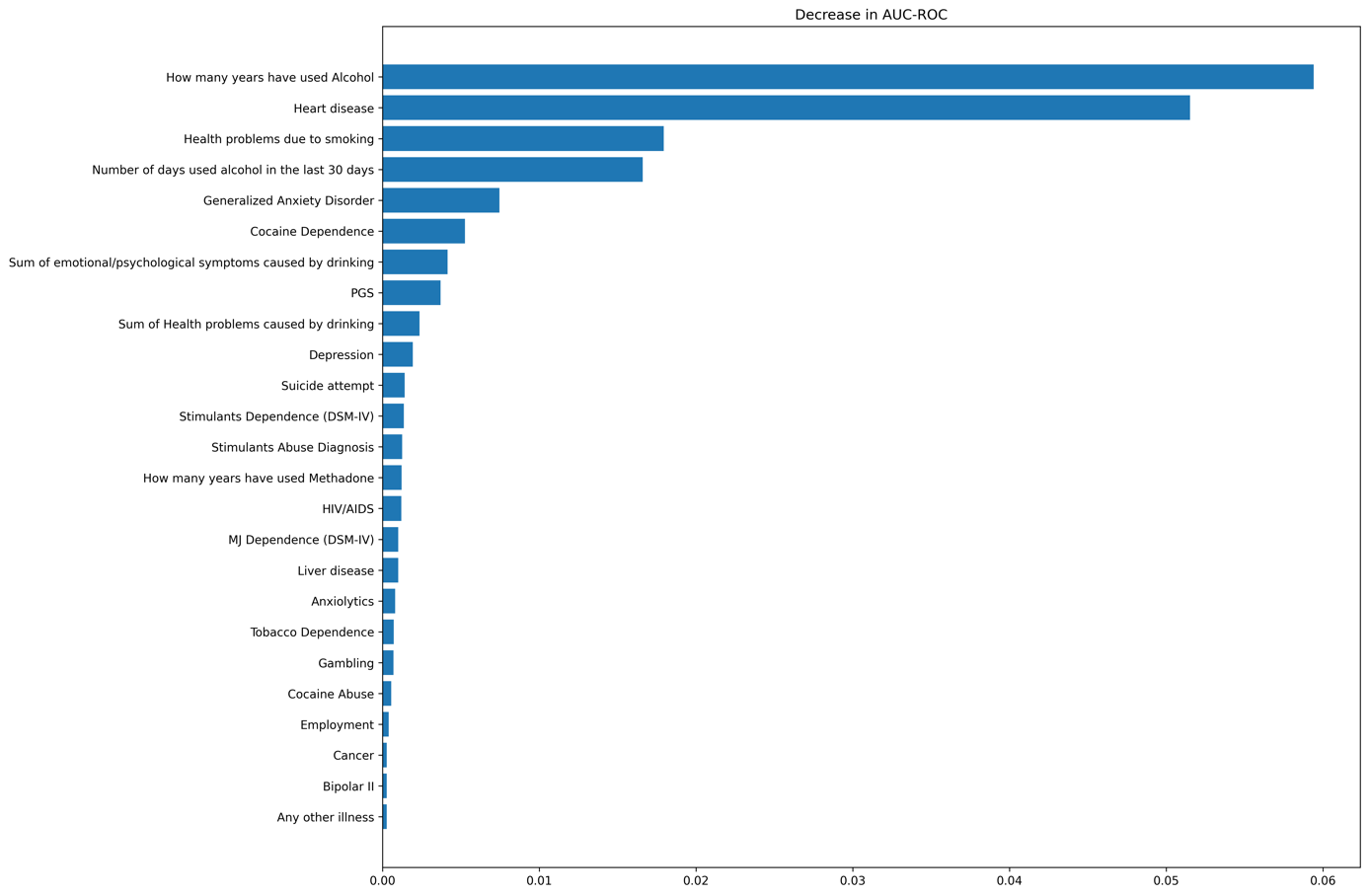

E) EUR-only with PGS, >= 40 years

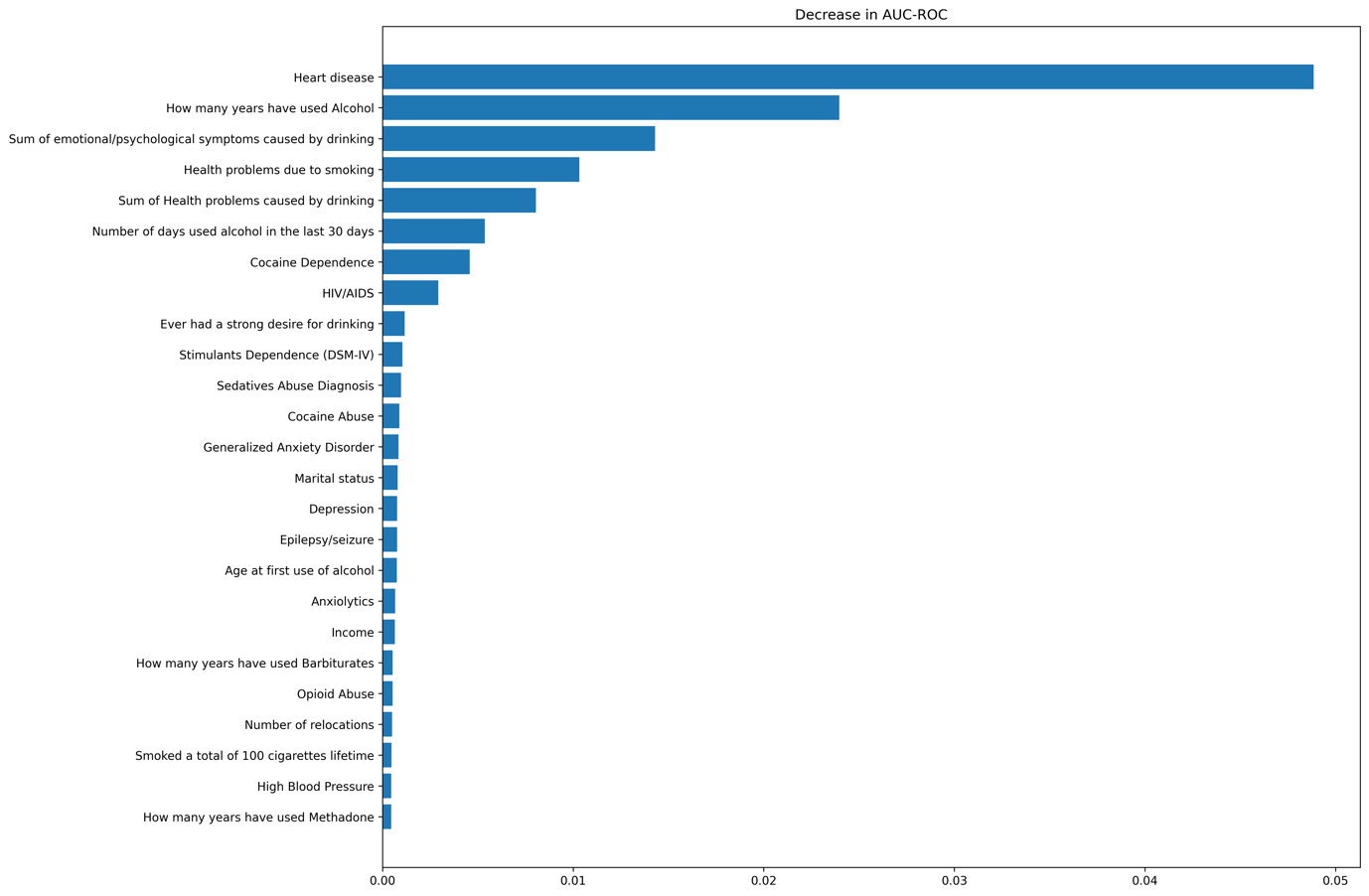

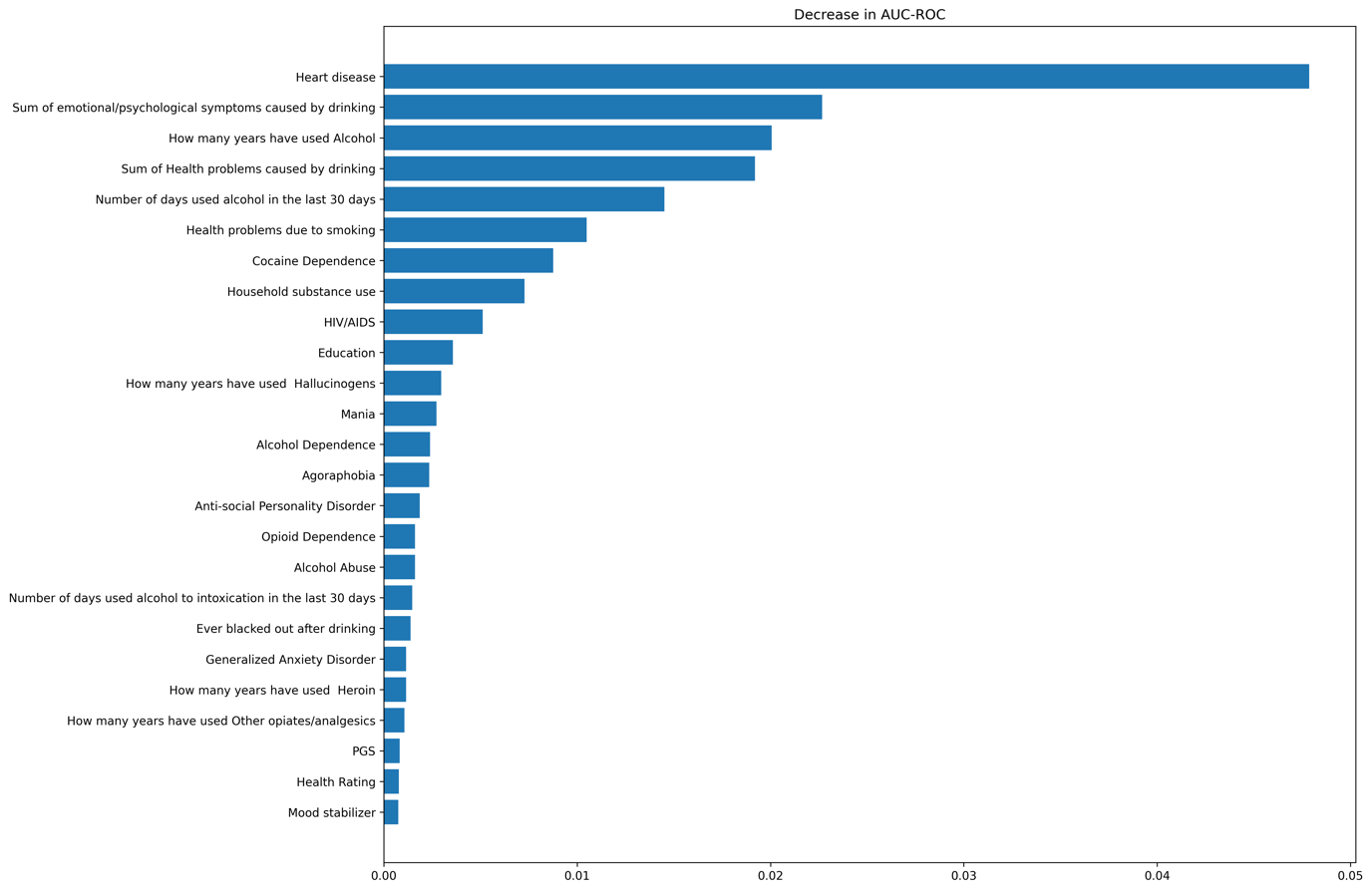

F) AFR-only without PGS

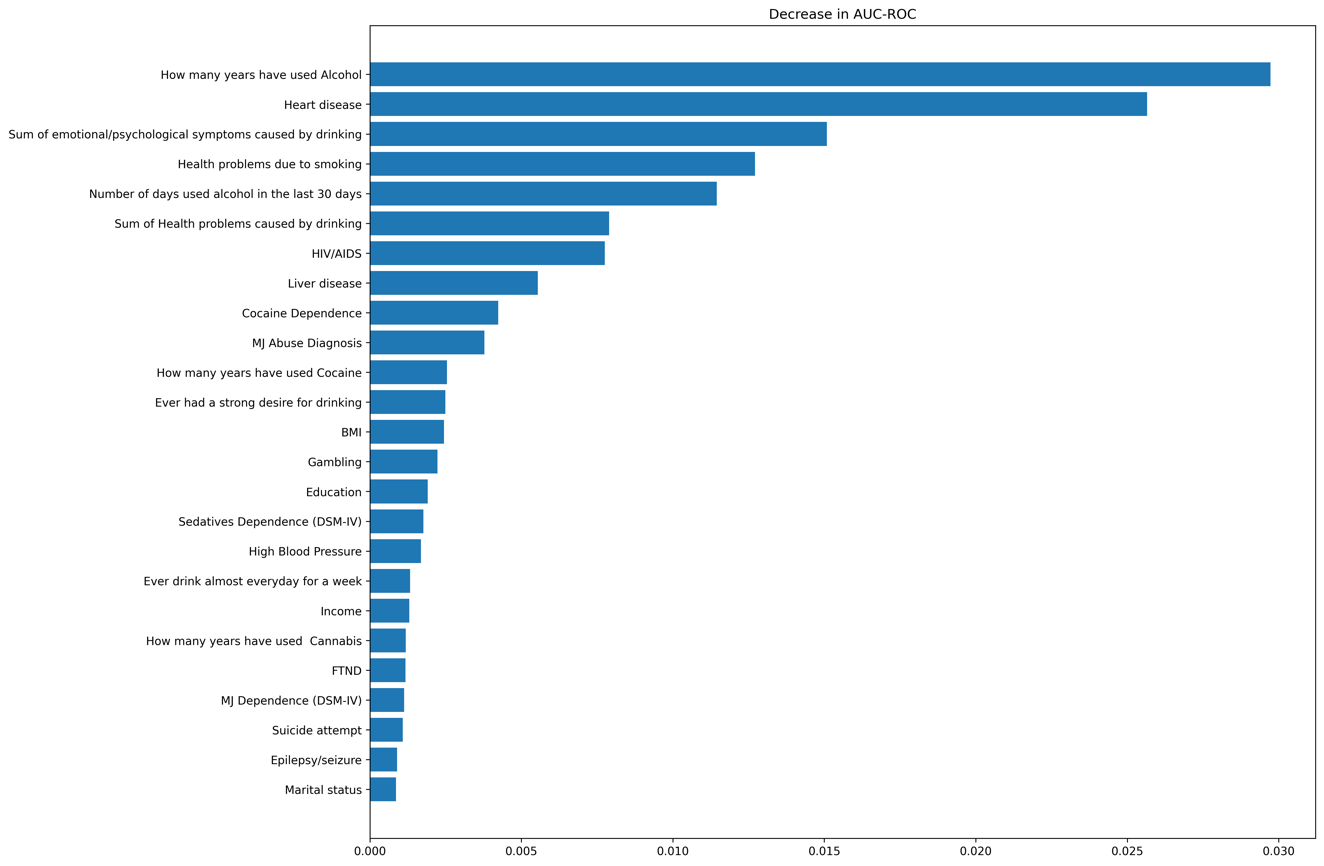

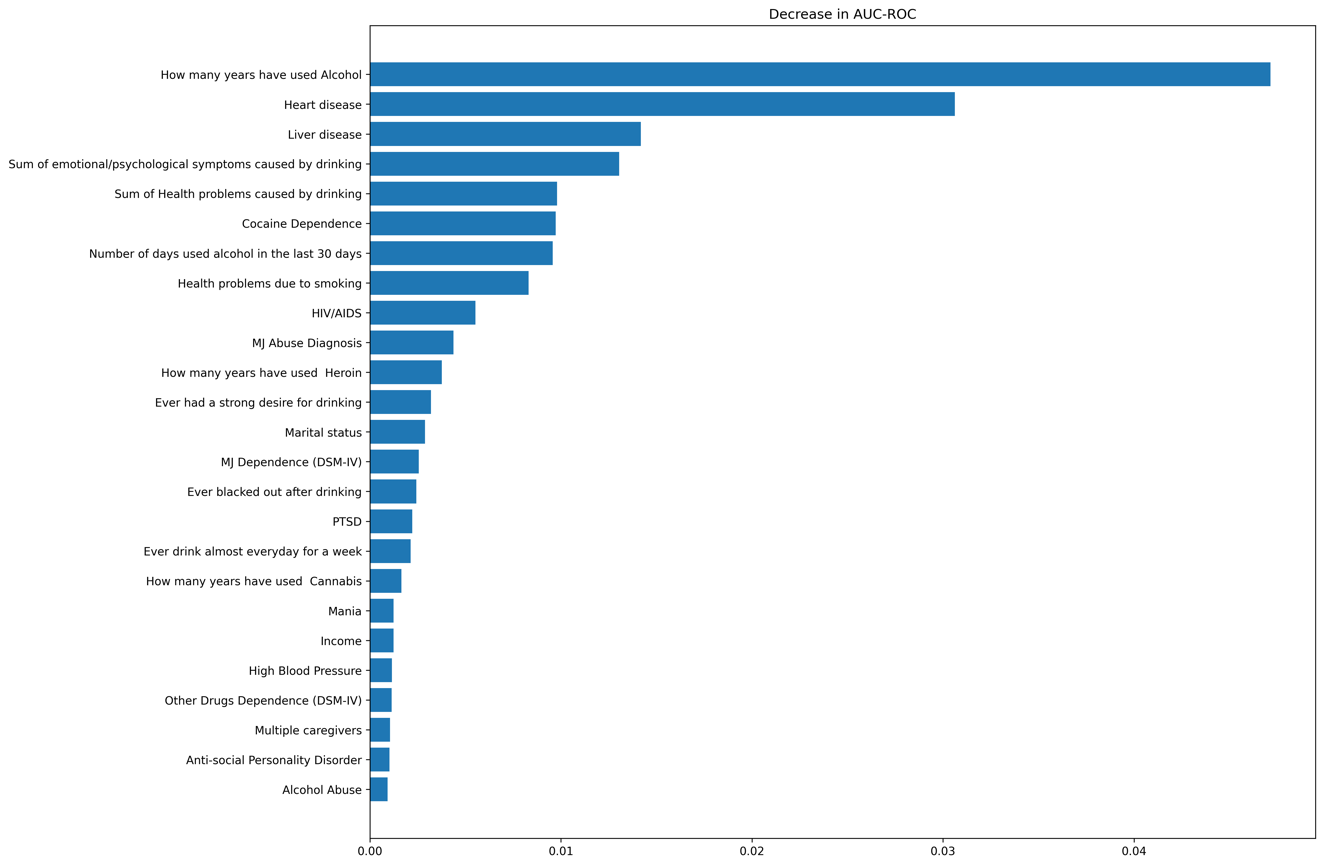

G) AFR-only with PGS

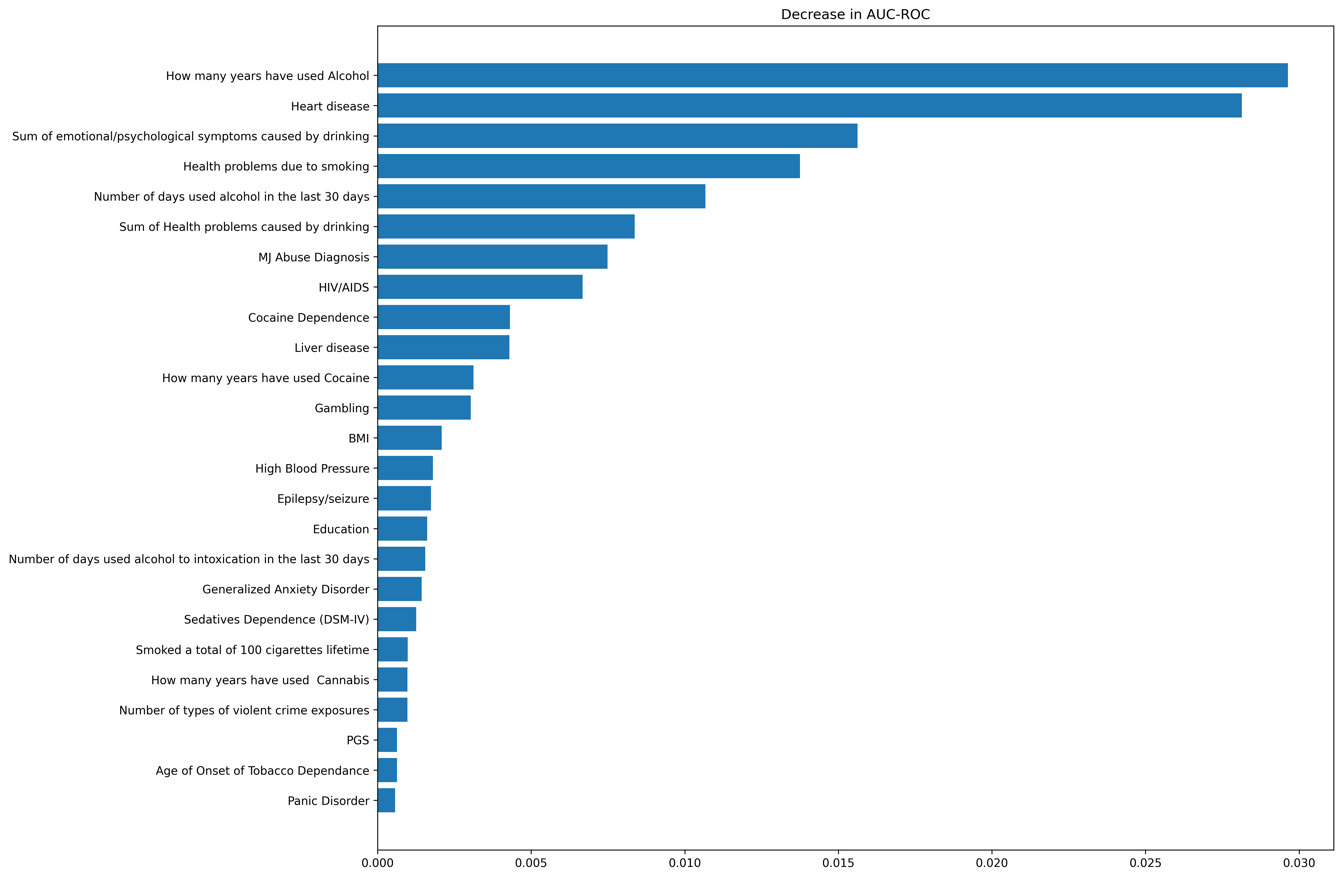

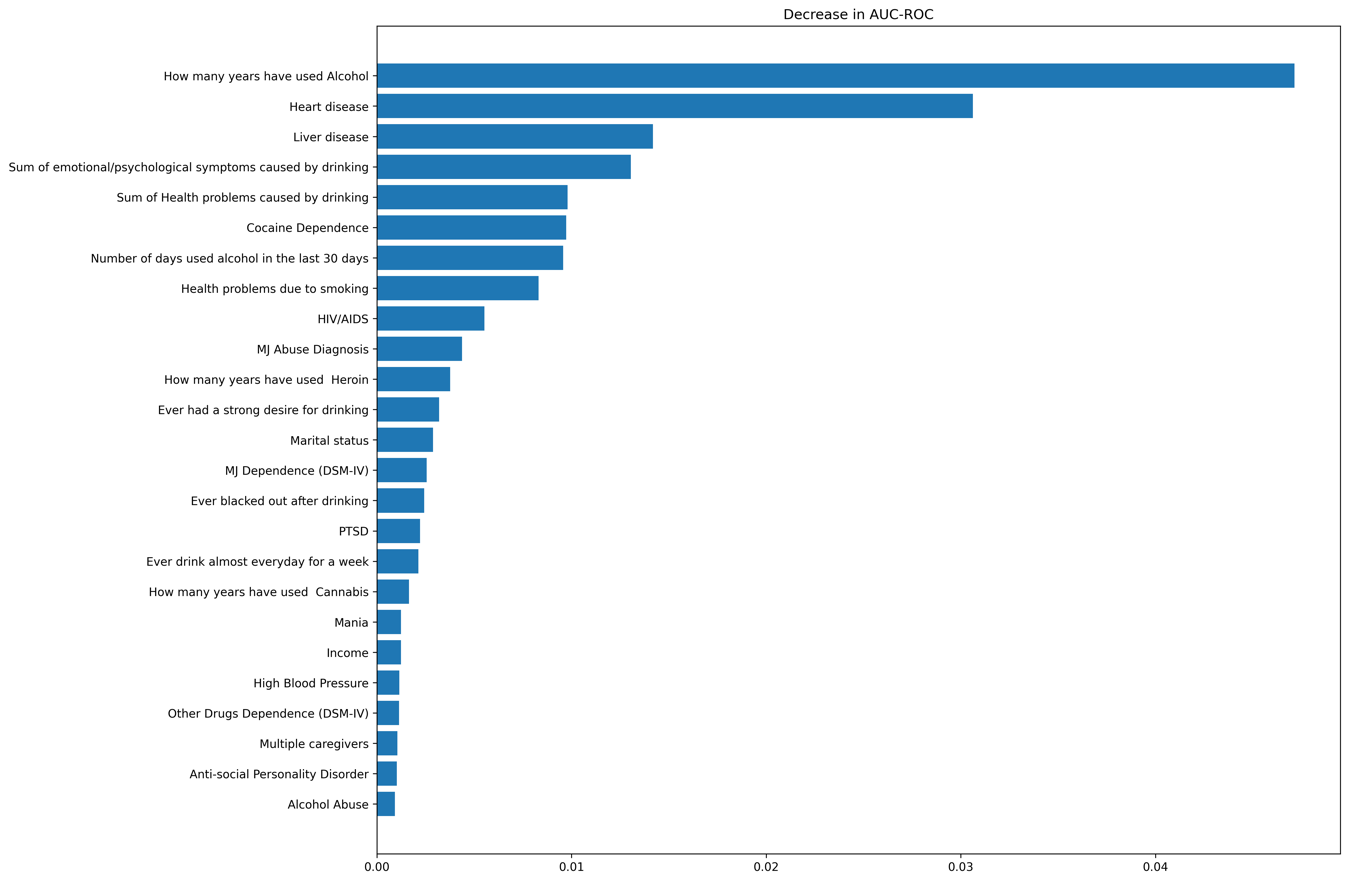

H) AFR-only with PGS, < 40 years

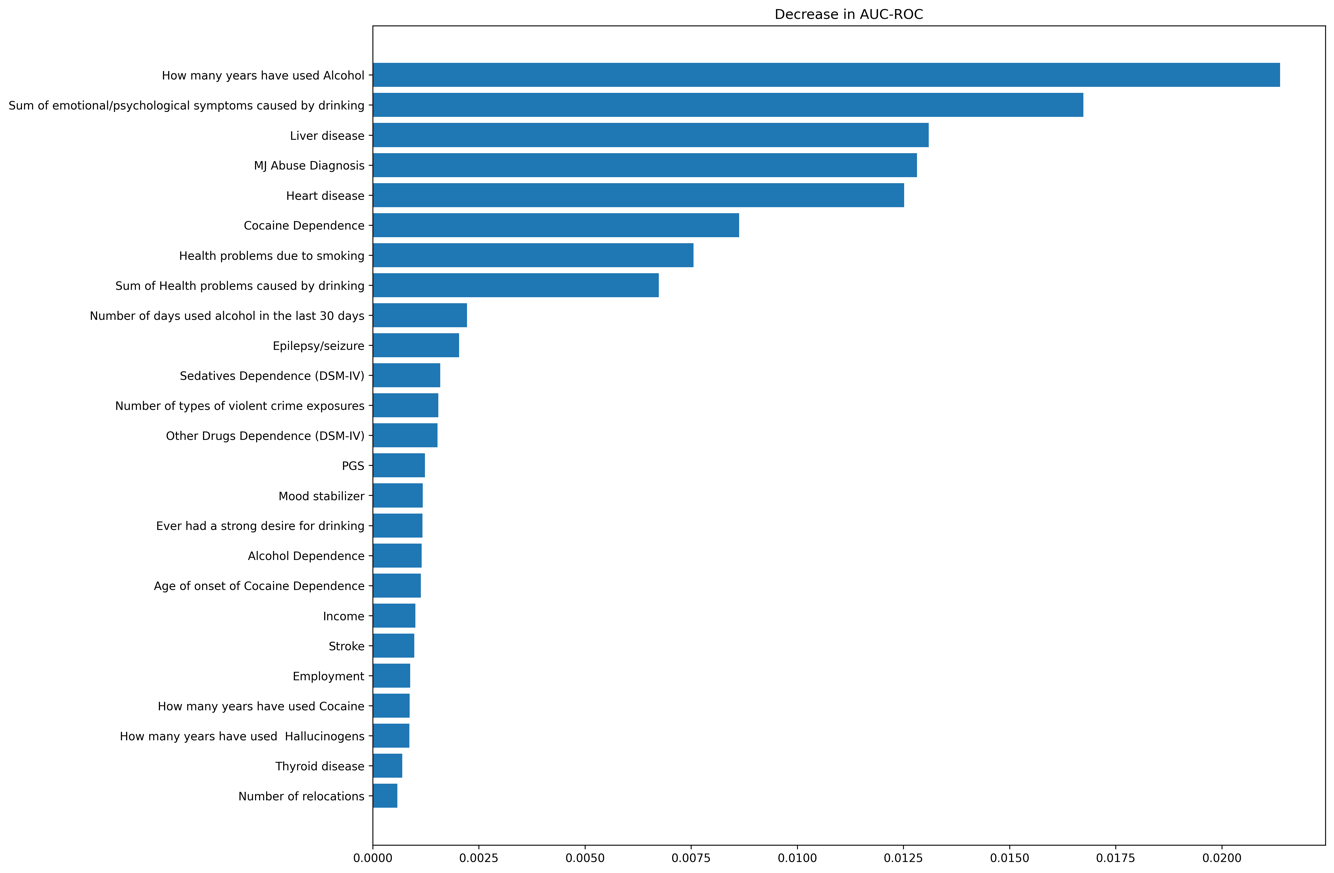

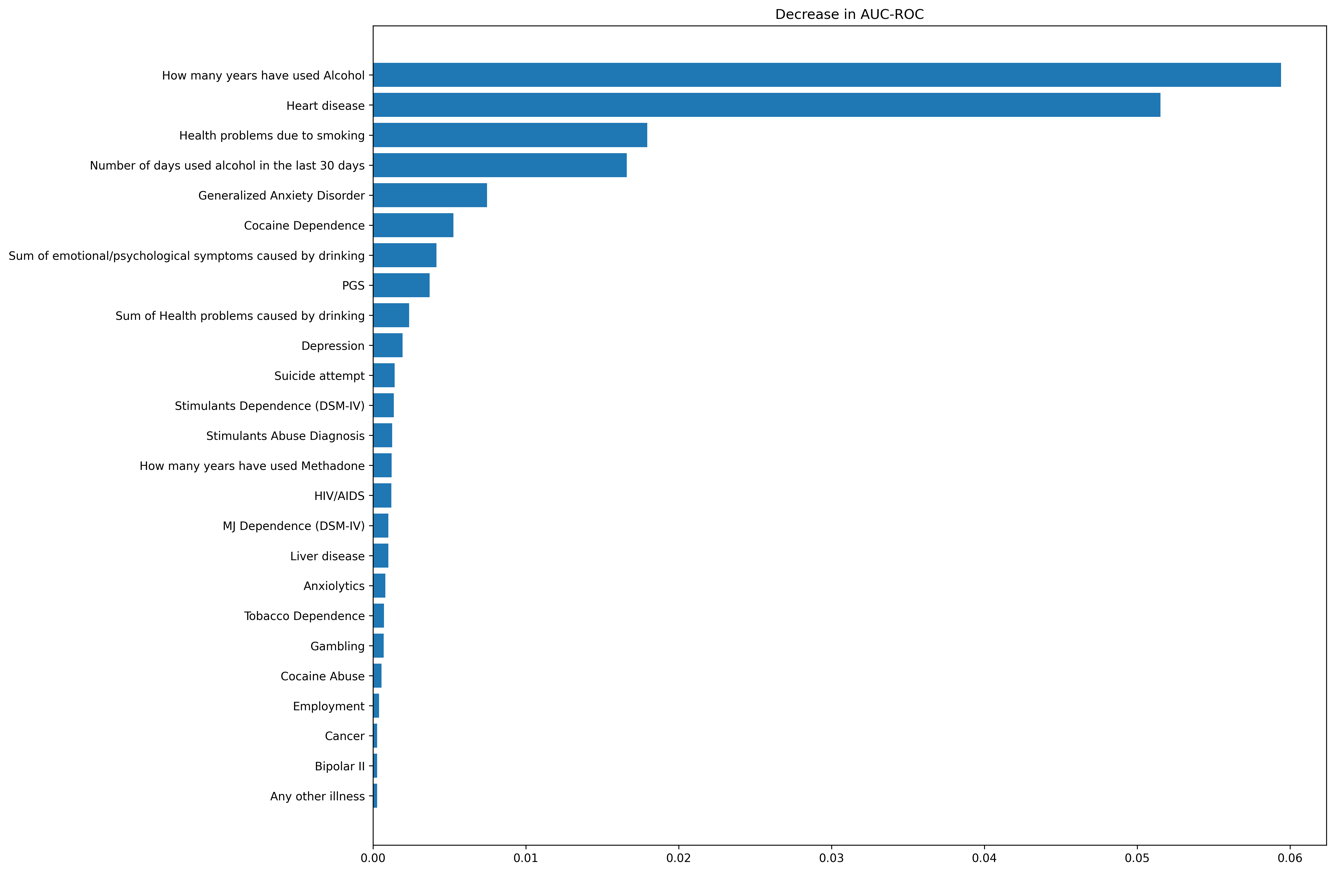

I) AFR-only with PGS, >= 40 years

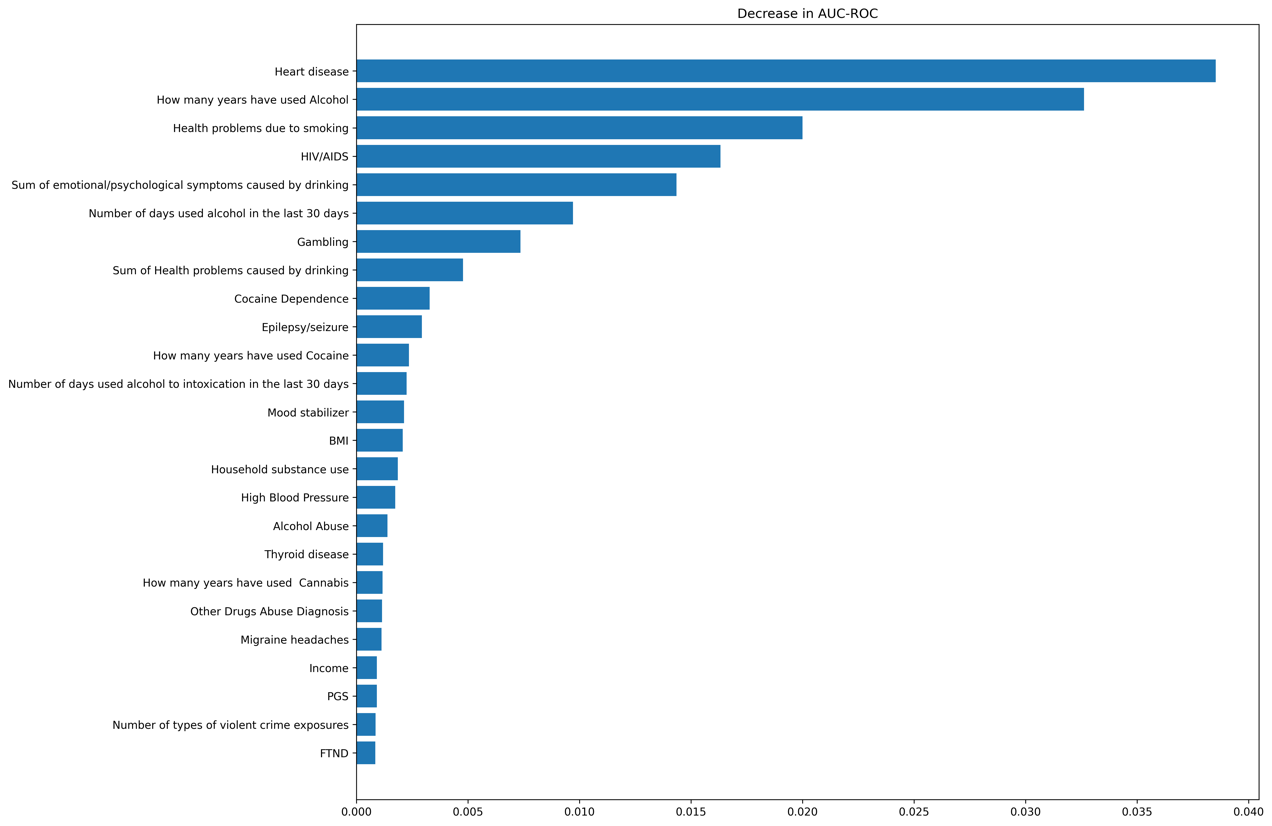

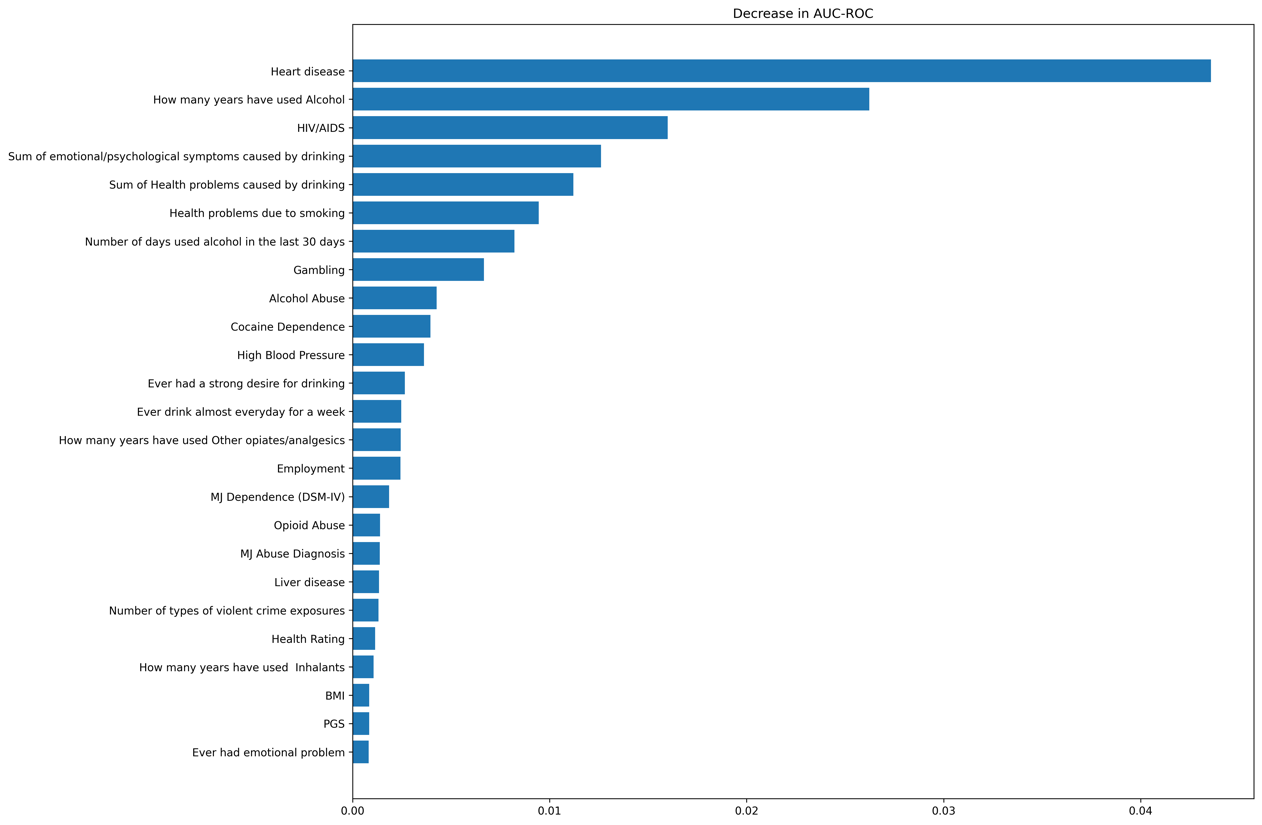
